# Simulation and prediction of spread of COVID-19 in The Republic of Serbia by SEIRDS model of disease transmission

**DOI:** 10.1101/2020.10.21.20216986

**Authors:** Slavoljub Stanojevic, Mirza Ponjavic, Slobodan Stanojevic, Aleksandar Stevanovic, Sonja Radojicic

**Author notes:** Corresponding author: Slavoljub Stanojevic, e mail.

## Abstract

As a response to the pandemic caused by SARS-Cov-2 virus, on 15 March, 2020, the Republic of Serbia introduced comprehensive anti-epidemic measures to curb COVID-19. After a slowdown in the epidemic, on 6 May, 2020, the regulatory authorities decided to relax the implemented measures. However, the epidemiological situation soon worsened again. As of 7 February, 2021, a total of 406,352 cases of SARSCov-2 infection have been reported in Serbia, 4,112 deaths caused by COVID-19. In order to better understand the epidemic dynamics and predict possible outcomes, we have developed an adaptive mathematical model SEAIHRDS (S-susceptible, E-exposed, A-asymptomatic, I-infected, H-hospitalized, R-recovered, D-dead due to COVID-19 infection, S-susceptible). The model can be used to simulate various scenarios of the implemented intervention measures and calculate possible epidemic outcomes, including the necessary hospital capacities. Considering promising results regarding the development of a vaccine against COVID-19, the model is extended to simulate vaccination among different population strata. The findings from various simulation scenarios have shown that, with implementation of strict measures of contact reduction, it is possible to control COVID-19 and reduce number of deaths. The findings also show that limiting effective contacts within the most susceptible population strata merits a special attention. However, the findings also show that the disease has a potential to remain in the population for a long time, likely with a seasonal pattern. If a vaccine, with efficacy equal or higher than 65%, becomes available it could help to significantly slow down or completely stop circulation of the virus in human population.

The effects of vaccination depend primarily on: 1. Efficacy of available vaccine(s), 2. Prioritization of the population categories for vaccination, and 3. Overall vaccination coverage of the population, assuming that the vaccine(s) develop solid immunity in vaccinated individuals. With expected basic reproduction number of Ro=2.46 and vaccine efficacy of 68%, an 87% coverage would be sufficient to stop the virus circulation.

## 1. Introduction

On 11 March, 2020, the World Health Organization characterised the disease caused by the novel SARS-Cov-2 virus as a pandemic [1]. The Initial epidemic outbreak in China spread outside the Wuhan area, and subsequently on a global scale. On 6 March, 2020, the first case of the novel coronavirus infection was reported in the Republic of Serbia. Taking into consideration the escalation of the disease and limited effects of the initially implemented measures, the state of emergency was declared throughout the country on 15 March, 2020. Comprehensive anti-epidemic measures (e.g. lockdown of entire country) were introduced in the entire country [2].

Due to the absence of specific pharmaceutical intervention, Serbia, like other countries, implemented an anti-epidemic strategy based on physical distancing, school and university closures, reduced number of workers present in the workplaces, closure of places of worship for public religious services, reduced working hours of cafés and restaurants, avoiding mass gatherings, events, sports games, tracing and identification of infected people’s contacts, etc. After a slowdown in the epidemic, as shown in the relevant officially published data, the regulatory authorities decided to relax the introduced measures on 6 May, 2020. However, the epidemiological situation soon worsened again, resulting in the reinstatement of some measures, as well as the introduction of new measures [2]. Although the return of extensive measures has yielded favourable results, the further development of the epidemic is not clear.

For these reasons, mathematical modelling has a crucial role in understanding the epidemic and predicting possible outcomes. Modelling is a particularly useful tool for devising strategies for combating the epidemic, capacity planning, and selection of efficient measures, especially in the absence of specific pharmaceutical treatments [3, 4, 5]. Mathematical modelling based on differential equations dates back to the first half of 20^th^ century. In 1927, Kermack and McKendrick developed the basic model of disease transmission consisting of three compartments: susceptible (S), infected (I) and removed (R). The model was based on a connected system of nonlinear differential equations as a special case of the general epidemiological model [6, 7]. Subsequent models, became more complex and adapted to the needs of research [5].

Since the outbreak of COVID-19, many published papers have dealt with the implementation of mathematical modelling and prediction of possible outcomes of COVID-19 epidemics. Most of these research efforts have been based on the implementation of the SIR (susceptible-infected-removed) model. Many of the other models provide a clear picture of dynamics of COVID-19 spreading, including the overloading of the relevant health systems. For example, Ferguson et al., developed one of the first models for COVID19 simulation, which was, among other things, used to plan the health care resources [8]. Wu et al., developed the SEIR model to examine the dynamics of SARS-Cov-2 transmission from person to person. This model was also used to calculate the basic reproduction number *R*_*o*_, which we use in this paper as one of the key parameters [9]. The classical SIR model assumes that there is homogenous mixing of infected and susceptible populations and that the total population is constant and does not change over time. Moreover, according to the classical SIR model, there is a monotonous decline in susceptible population towards zero [10]. However, such assumptions are not objective in the case of COVID-19 spreading and they are the basic problem in the modelling of this pandemic. In reality, the human population fluctuates constantly [11]. In order to account for such fluctuation, and better understand the COVID-19 epidemic in the Republic of Serbia, we have employed mathematical modelling of the epidemic using the available data on the characteristics of the disease, such as incubation period, latent period, recovery period, severity of clinical signs, and mortality rate caused by COVID-19.

Unlike the classic SIR model, the SEAIHRDS (S-susceptible, E-exposed, A-asymptomatic, I-infected, H-hospitalized, R-recovered, D-dead due to COVID-19 infection, S-susceptible) epidemic model, developed for this research, simulates the spreading of COVID-19 in an open population. Taking into account that the population is constantly changing and that various measures are applied for different strata or subgroups of the population (such as preschool children, children attending primary school, high school students, students, employees, the unemployed and retirees), as well as changes in the intensity of applied measures, we have proposed the use of a model that takes these circumstances into account. Based on input disease parameters taken from scientific literature and specific data related to Serbia, this model simulates daily disease occurrence, including the number of hospitalized patients and cases which require intensive care. The model also predicts the expected number of deaths, as well as hospital capacities necessary to accommodate the patients. It provides a possibility to simulate different scenarios of disease control and intervention measures. Considering the expectations of successful development of the vaccine against COVID-19 in the near future, we added an option to model vaccination of different strata of the population as a set of disease control strategies.

## 2. Methodology

This section presents the research methodology and the proposed model, which were used to predict the further dynamics of the epidemic in Serbia. We also presented the data that were used to model the epidemic, a simulated strategy to combat COVID-19, and a sensitivity analysis.

### 2.1. SEAIHRDS mathematical model

Classical SEIRDS model divides the population into compartments, i.e. groups, and follows the disease dynamics at all times. The population is divided into the following compartments: the portion of the population susceptible to the infection is denoted by S, those latently infected with SARSCov-2 (exposed to) are denoted by E, the infected individuals who are able to spread the disease are denoted by I, the ones recovered from the infection are denoted by R, and those who died due to disease with D. Assuming that individuals mix homogenously, the force of infection *λ* (the rate at which susceptible persons are infected) is related to *per capita* contact rate *β*. Also, the risk of infection is closely related to the number of infectious individuals in the population I_*t*_. It depends on the number of infectious individuals (I_*t*_) and how frequently they make contacts with other persons. In a situation of homogenous mixing among the population, the force of infection λ can be express as follows:

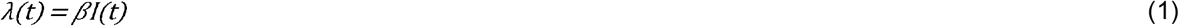

The change of rates in every compartment per unit time in SEIRDS model is presented in the following series of differential equations:

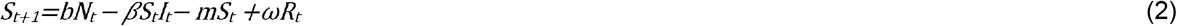

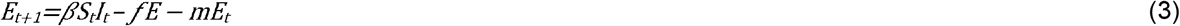

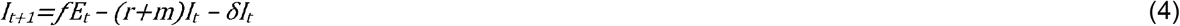

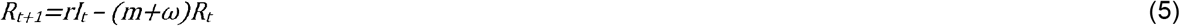

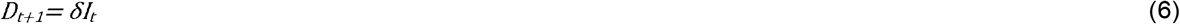

where *f* is rate of onset of infectiousness expressed as the reciprocal of the latent infection period, r is the rate at which infectious individuals are recovered, *δ* is the rate at which infectious individuals die from COVID-19 infection and ω is rate of waning of immunity. The total population at any particular interval of time *t* is:

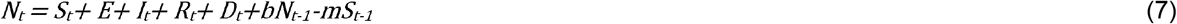

where parameters *b* and *m* are *per capita* daily birth rate and death rate unrelated to COVID-19.

However, considering that implemented anti-epidemic measures against COVID-19 do not have an identical impact on the population’s age subgroups and that COVID-19 pathogenesis varies in different age subgroups, we propose the use of multi-compartment version of standard SEIRDS model. The model, named SEAIHRDS, monitors the dynamics of following compartments: susceptible individuals (*S*), latently infected with SARSCov-2 (*E*) (exposed to/presymptomatic), asymptomatic infectious individuals (*A)*, infectious individuals with symptoms/clinically ill (*I*), hospitalized patients (*H*), recovered individuals (*R*), and those who died due to disease (*D)*. In this model the susceptible population was further stratified within the compartment S according to age and occupations. Grouping into various strata was done according to the real age structure of the Republic of Serbia population as follows: pre-school children (*S*_*ps*_), elementary school children (*S*_*es*_), high school children (*S*_*hs*_), college students (*S*_*cs*_), unemployed population (*S*_*ua*_), employed population (*S*_*ea*_), and elderly/retired (*S*_*r*_) (Table 1). To simulate the epidemic progression through different population strata-subgroups, we used appropriate, stratum-specific, model parameters and factors of effective contact reduction (anti-epidemic intervention measures -*ρ*), which were adapted to the relevant population groups: lockdown of the entire country, closures of pre-school, school and college sessions closures, reduced number of workers allowed in the workplaces, work from home, restrictions of mobility of elderly people, etc. During the simulation, we monitored the effects of various levels of contact reduction, ranging from 20% to 75%, taking into account the realistic possibilities of maintaining a minimum work process, functioning of the society and feasibility of such measures.

**Table 1.**
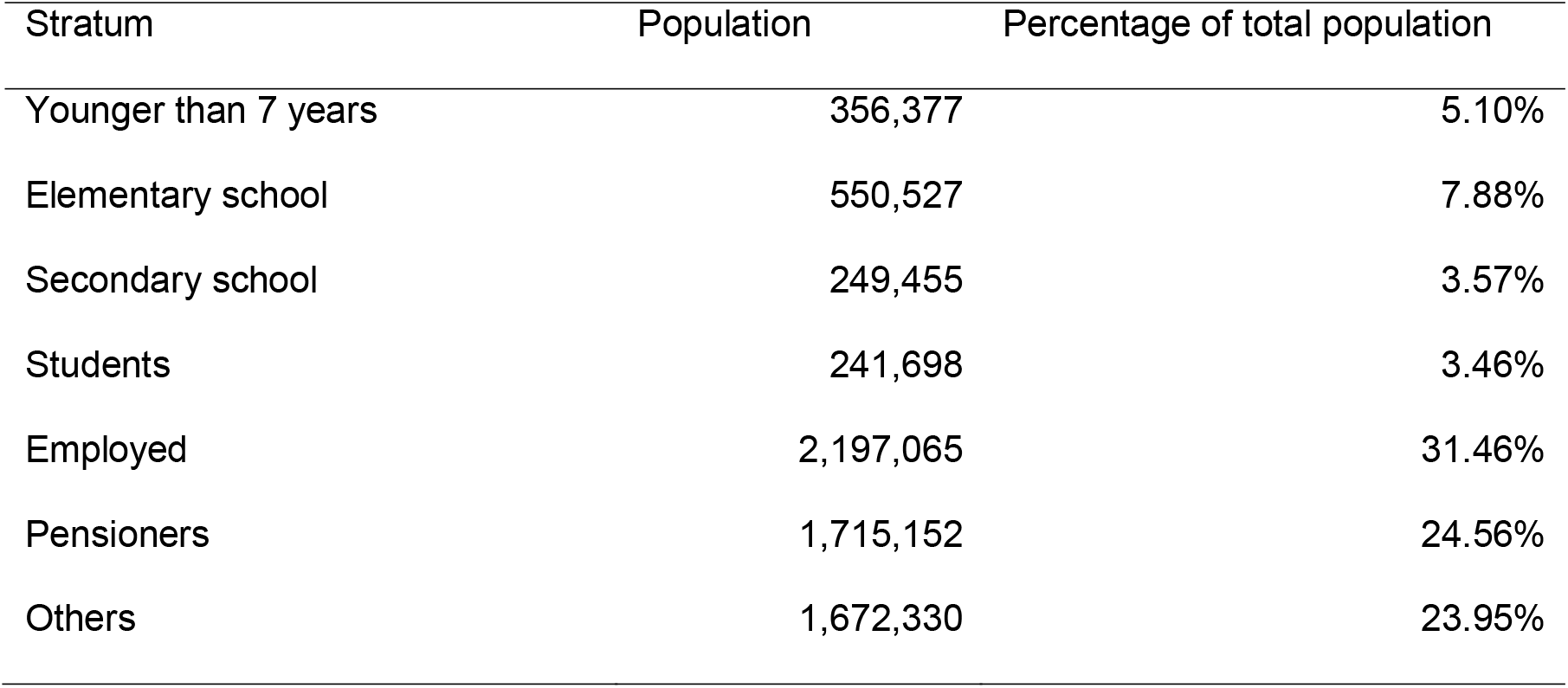
Structure of different population strata in the Republic of Serbia [23]

Given that intervention measures, applied in response to the emergence of COVID-19, are not the same for all population strata, homogeneous mixing can be expected only within same population stratum. The rate of effective contacts *β*, after the application of intervention measures, is no longer identical at the level of all strata of the population. Effective contacts are limited by the intensity and types of measures applied and are identical only when it comes to individuals within the same population strata. Furthermore, persons in different population strata become infected at different rates depending on how frequently they interact with other persons in their own subgroup and other subgroups. If we assume that force of infection differs between different strata of population, the equation for the force of infection would be as follows:

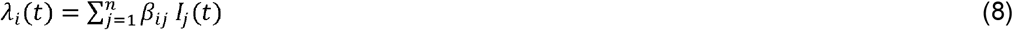

where *λ*_*i*_(*t*) is force of infection in the *i*^*th*^ population strata, *β*_*ij*_ is the rate at which susceptible persons in the *i*^*th*^ population strata and infectious persons in *j*^*th*^ population strata come into effective contact per unit of time, *and I*_*j*_(*t*) is the number of infectious persons in *j*^*th*^ population strata. Also, in this model the number of recovered and dead is conditioned with different ages and genders.

Now our model will be expressed as follows:

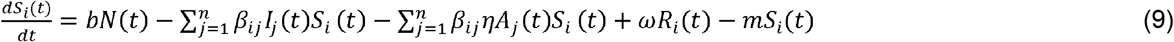

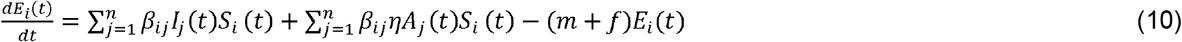

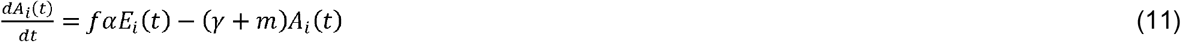

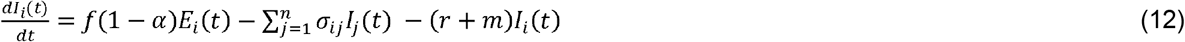

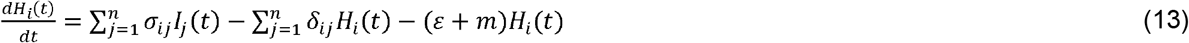

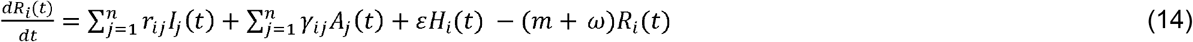

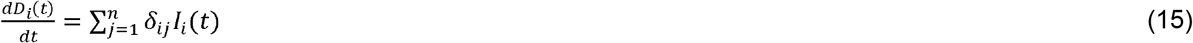

where *α* is the proportion of asymptomatic cases, *η* accounts for the relative infectiousness of asymptomatic carriers (in comparison to symptomatic carriers), *r* is the rate at which infectious individuals whit symptoms are recovered, *γ* is the rate at which asymptomatic infectious individuals are recovered, a is rate at which infectious individuals are hospitalized, *ε* is the rate at which hospitalized patients are recovered, δ is the rate at which infected individuals die from COVID-19 infection and ω is rate of waning of immunity (Supplementary material).

### 2.2. Determining the herd immunity threshold and control of COVID-19 by vaccination policy

Considering the undergoing worldwide efforts to develop a vaccine against COVID-19 and promising results, we extended the model to simulate and analysed the effects of a hypothetical vaccination on the epidemic dynamics, and to estimate the extent of coverage of vaccination which can interrupt the chain of infection. The control of COVID-19 by vaccination means targeting the entire susceptible population with mass vaccination until critical herd immunity achieved. In such situation there is a “race” between the exponential growth of epidemic and mass vaccination. The level of herd immunity threshold (HIT) can be calculated by the following formula:

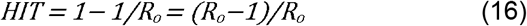

and the critical vaccination coverage required to achieve herd immunity can be obtained by multiplying herd immunity threshold with reciprocal value of vaccine efficacy, *v*_*e*_:

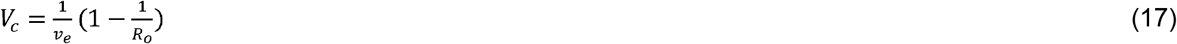

Most people infected with SARS-CoV-2 develop an immune response followed by the development of specific antibodies between 10 and 21 days after getting infected [12]. Specific IgM and IgG antibodies against SARS-CoV-2 develop 6 to 15 days after the onset of the disease [13-17]. According to some studies, the presence of antibodies has been confirmed in less than 40% of the patients within 1 week after the onset of the disease, whereas percentage reaches 100% of subjects 15 days after the onset of disease [18]. Although duration of the immune response against CVOVID 19 is still unknown, comparing with other coronaviruses, where immunity wane within 12 to 52 weeks after the first symptoms appear [19, 20], while in the case of SARS-CoV-1 infection the presence of IgG antibodies was confirmed in 90% and 50% of infected patients, respectively, over two and three years, respectively [29], we assumed that durable immunity against COVID-19 is possible [20, 21]. Immunity to HCoVOC43 and HCoV-HKU1 appears to wane appreciably within 1 year [21, 22], whereas SARS-CoV-1 infection can induce longer-lasting immunity [23]. S. F. Lumley et. in a study conducted on 1,246 persons recovered from COVID-19 found no symptomatic re-infections over 6 months [24].

Based on these findings, and fact that SARS-Cov-2 virus is also beta coronavirus, we assumed that in the event of the development of a successful vaccine, immunity against the SARS-Cov-2 virus could last for a year, as well as after recovery after a natural infection. In a study conducted to determine the dynamics of SARS-Cov-2 transmission in the post pandemic period, Kissler et al. applied a similar approach in defining the possible length of the immunity period [20].

For the purpose of modelling a control strategy based on vaccination, additional compartment to the model was added denoted with V(t), in which there are vaccinated persons who have successfully developed protective immunity after vaccination. The vaccination parameter, υ, is the daily rate of vaccination of susceptible population and it represents the proportion of susceptible population immunized per unite time. The critical daily rate of vaccination, *⍰*_c_, is *⍰*_*c*_= (*b*+*ω*)(*R*_*o*_ −1), required to interrupt the infection [5]. The basic reproduction number under the vaccination is *R*_*op*_ *= (1-p)R*_*o*_. The proportion of effectively protected persons, *p*, is conditioned by parameter the vaccine efficacy, *v*_*e*_. This parameter represents a proportion of person who successfully developed protective immunity after vaccination, whereas total number of actively protected individuals in time t is V(t) = number of vaccinated x *v*_*e*_ [5]. In this compartment the daily rate of waning of immunity at which immunity of vaccinated population fades out is ω, and it is reciprocal to the period of lasting of immunity. Vaccinated persons, after losing their immunity, become sensitive again and removed to the compartment S. The change of rate in this compartment per unit time is as follows:

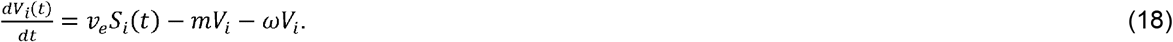

The compartment S(t) is slightly modified as follows:

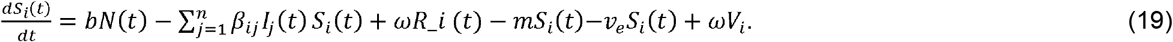

The other of compartments of SERIDS model remain unchanged.

### 2.3. Model parametrisation

In proposed model, *β* is *per capita* effective contact rate at which specific persons come into effective contact per unit of time. An effective contact is defined as a contact sufficient to cause disease transmission [10, 24, 26]. We calculated the parameter *β* by using the formula:

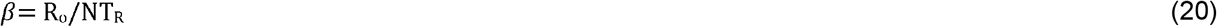

where R_o_ is a basic reproduction number of the disease, i.e. the average number of newly infected people with COVID-19 (secondary infection cases), infected by one infectious individual in a totally susceptible population, N is total population, and T_R_ is the average duration of infectious period [10, 25, 26]. The R values of 2.46 and 3.1 are adopted from the relevant literature. The R_o_ values were based on the data obtained during the initial phase of the epidemic in Italy [27]. Since the implemented measures and disease transmission were simulated through various population strata, we corrected the {3 parameter with a relevant, stratum-specific contact reduction factor ρ_*i*_. In this way, we obtained the *per capita* contact rate specific for each separate stratum based on following formula

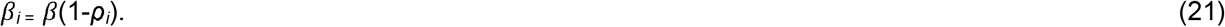

The values of the *ρ*_*i*_ factor in different population strata ranged from 0.25 to 0.75 (effective contact reduction ranged from 25% to 75%).

Parameters such as daily birth and death rates were calculated based on the data published by the Office of Statistics of the Republic of Serbia, and data published by the World Bank regarding the life expectancy in the Republic of Serbia [28, 29]. The latter study reported that the life expectancy in Serbia was 79.06 years in 2017 [29]. By using this figure, we expressed the daily death rate as a value reciprocal to life expectancy *m* = 0.000036. We calculated the daily *per capita* birth rate of *b= 0*.*000025* based on the figure of 9.2 births in the Republic of Serbia per 1000 people in a year. These estimates were needed to realistically simulate fluctuations of the total population. To simplify the calculations, we assumed that the general morality rate *m* is applicable for all population strata.

The infectivity rate, i.e. the rate of transfer from compartment E to I, was derived as a value reciprocal to the COVID-19’s average latent period. The data on the average duration of latent infection (*f*^-1^) and the average period during which an infected person is shading the SARS-Cov-2 virus (*T*_*R*_) were adopted from the relevant literature as *f*^-1^ = 3.5 days [30, 31] and *T*_*R*_ = 9.3 days, respectively [30]. Also the data on the percentage of hospitalized patients and those whose therapy requires intensive care, used for prediction of required hospital capacities, as shown in Table 1, were taken from literature [30, 8].

Parameters such as *δ* and r are related to the infectious fatality rate (IFR) for COVID-19 and average times taken from onset of symptoms to death (*T*_*D*_) or recovery (*T*_*R*_). These parameters were calculated using the following formulas:

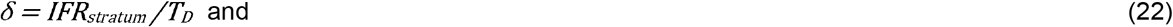

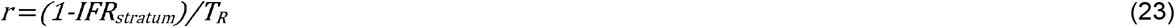

The IFRs, shown in Table 2, were taken from literature and compared with local IFR value which was calculated based on officially registered deaths published by the health system of the Republic of Serbia [2]. The Calculation of local IFR is presented in section 2.4. Population data, (e.g. total population, age structure, and stratification) are presented in Tables 1 and 2. A summary of all model parameters is given in Table 3.

**Table 2.**
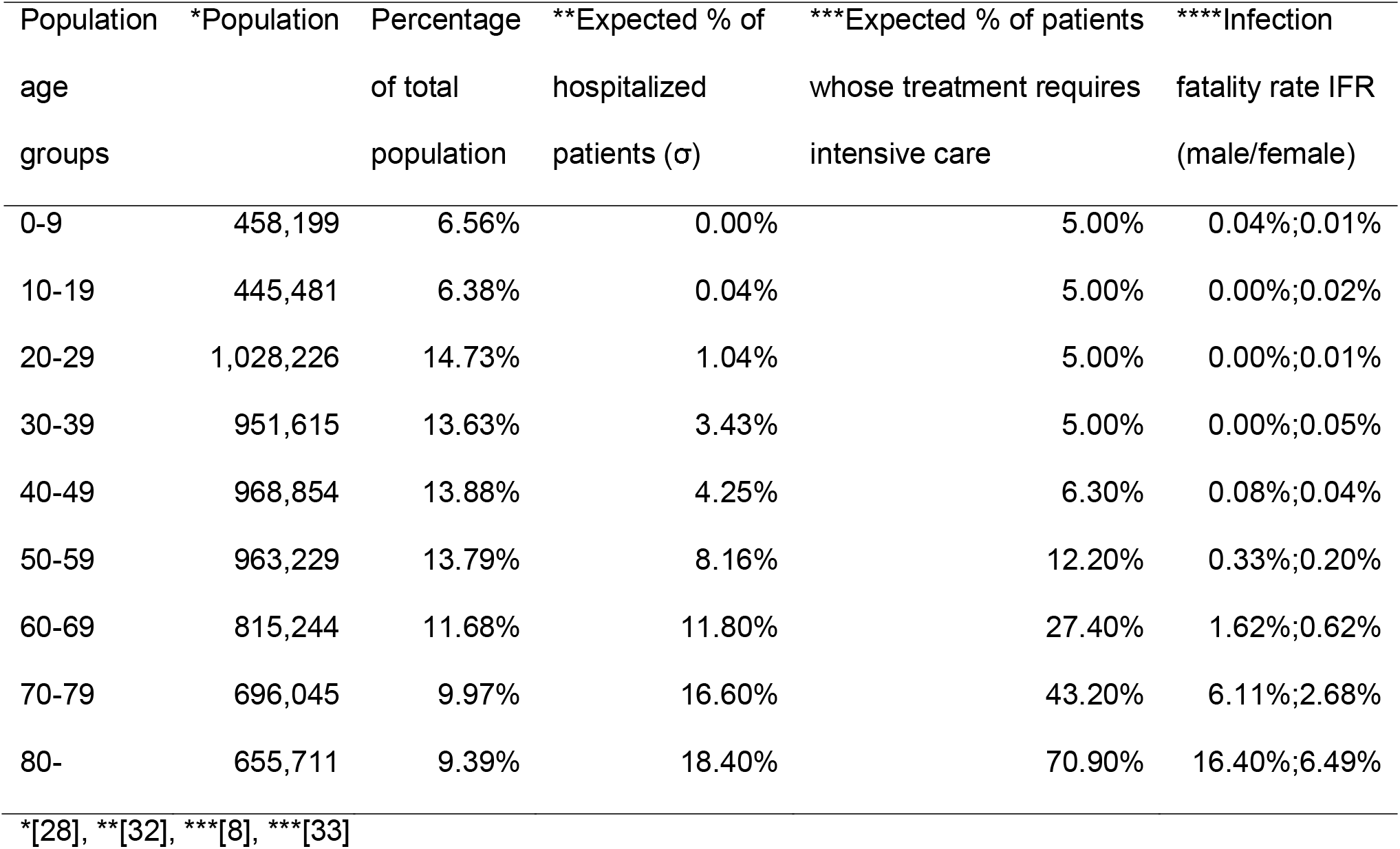
Age structure of the population of the Republic of Serbia and expected percentage of hospitalized patients, patients in intensive care, and death rate caused by COVID-19.

**Table 3.**
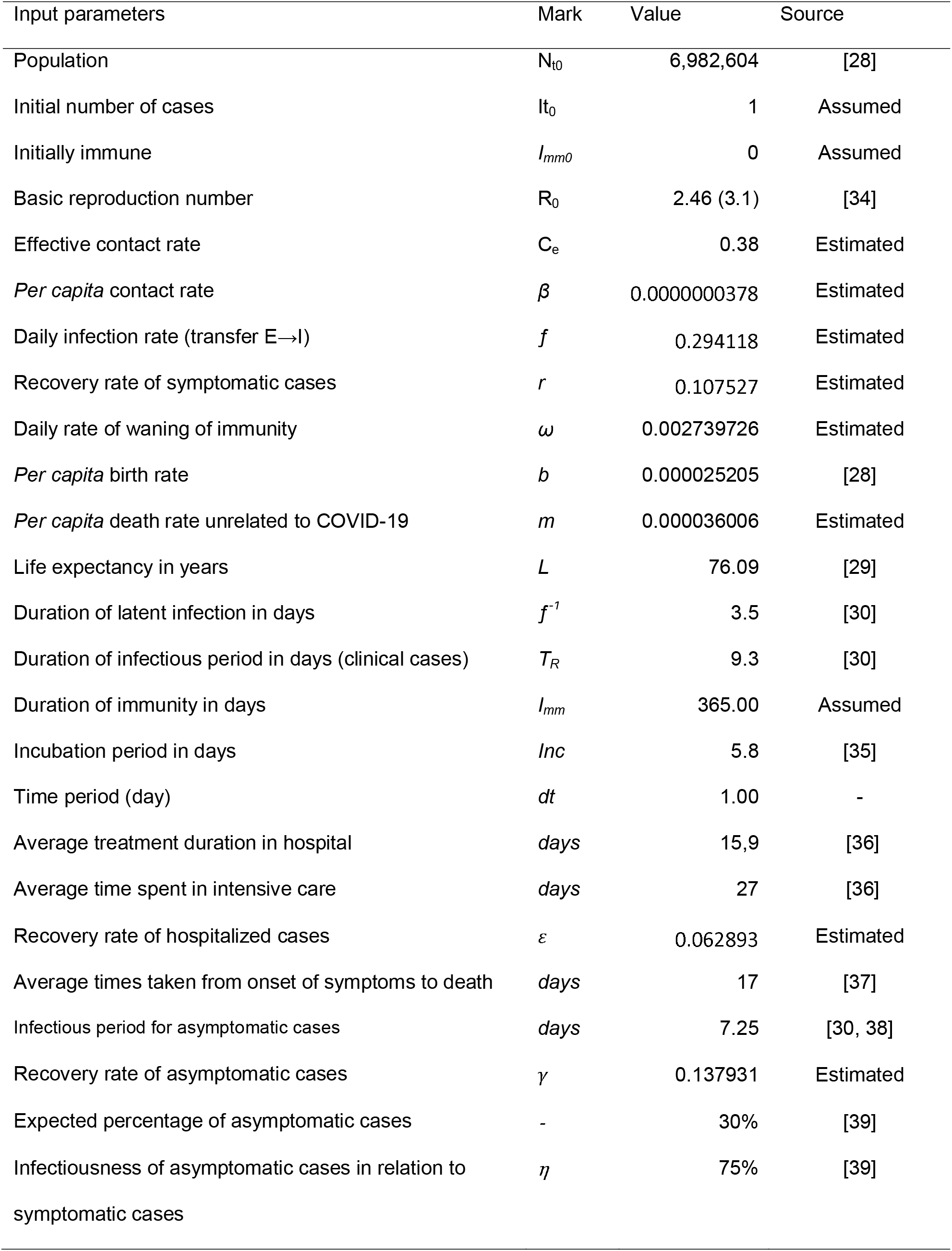
SEAIHRD model parameters

| Input parameters | Mark | Value | Source |
| --- | --- | --- | --- |
| Population | $N_{t0}$ | 6,982,604 | [28] |
| Initial number of cases | $I_{t0}$ | 1 | Assumed |
| Initially immune | $I_{mm0}$ | 0 | Assumed |
| Basic reproduction number | $R_0$ | 2.46 (3.1) | [34] |
| Effective contact rate | $C_e$ | 0.38 | Estimated |
| *Per capita* contact rate | $\beta$ | 0.0000000378 | Estimated |
| Daily infection rate (transfer E→I) | $f$ | 0.294118 | Estimated |
| Recovery rate of symptomatic cases | $r$ | 0.107527 | Estimated |
| Daily rate of waning of immunity | $\omega$ | 0.002739726 | Estimated |
| *Per capita* birth rate | $b$ | 0.000025205 | [28] |
| *Per capita* death rate unrelated to COVID-19 | $m$ | 0.000036006 | Estimated |
| Life expectancy in years | $L$ | 76.09 | [29] |
| Duration of latent infection in days | $f^{-1}$ | 3.5 | [30] |
| Duration of infectious period in days (clinical cases) | $T_R$ | 9.3 | [30] |
| Duration of immunity in days | $I_{mm}$ | 365.00 | Assumed |
| Incubation period in days | $Inc$ | 5.8 | [35] |
| Time period (day) | $dt$ | 1.00 | - |
| Average treatment duration in hospital | $days$ | 15,9 | [36] |
| Average time spent in intensive care | $days$ | 27 | [36] |
| Recovery rate of hospitalized cases | $\varepsilon$ | 0.062893 | Estimated |
| Average times taken from onset of symptoms to death | $days$ | 17 | [37] |
| Infectious period for asymptomatic cases | $days$ | 7.25 | [30, 38] |
| Recovery rate of asymptomatic cases | $\gamma$ | 0.137931 | Estimated |
| Expected percentage of asymptomatic cases | - | 30% | [39] |
| Infectiousness of asymptomatic cases in relation to symptomatic cases | $\eta$ | 75% | [39] |

### 2.4. Setting disease control scenarios

Five different scenarios were developed for simulating the COVID-19 epidemic control based on non-pharmaceutical interventions. SC1 implies a base-case scenario where the epidemic spreads in susceptible population without any anti-epidemic measures being implemented. In the other scenarios, the extent of contacts was reduced, for each population stratum separately, according to objective possibilities and measures which were implemented during the actual epidemic in the Republic of Serbia. Scenarios are presented in Table 4.

**Table 4.**
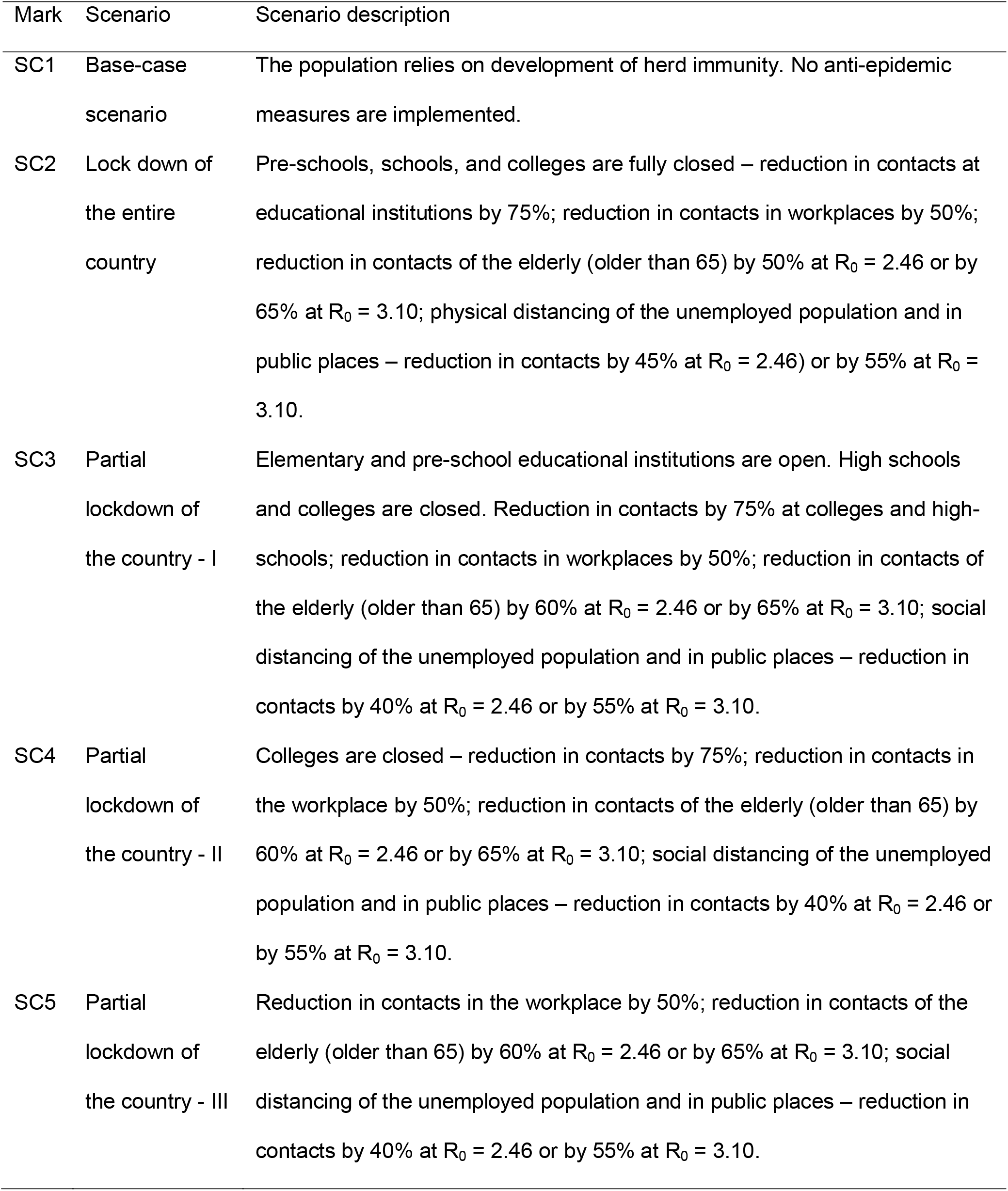
Description of different simulated non-pharmaceutical intervention scenarios

| Mark | Scenario | Scenario description |
| --- | --- | --- |
| SC1 | Base-case scenario | The population relies on development of herd immunity. No anti-epidemic measures are implemented. |
| SC2 | Lock down of the entire country | Pre-schools, schools, and colleges are fully closed – reduction in contacts at educational institutions by 75%; reduction in contacts in workplaces by 50%; reduction in contacts of the elderly (older than 65) by 50% at $R_0 = 2.46$ or by 65% at $R_0 = 3.10$; physical distancing of the unemployed population and in public places – reduction in contacts by 45% at $R_0 = 2.46$ or by 55% at $R_0 = 3.10$. |
| SC3 | Partial lockdown of the country - I | Elementary and pre-school educational institutions are open. High schools and colleges are closed. Reduction in contacts by 75% at colleges and high-schools; reduction in contacts in workplaces by 50%; reduction in contacts of the elderly (older than 65) by 60% at $R_0 = 2.46$ or by 65% at $R_0 = 3.10$; social distancing of the unemployed population and in public places – reduction in contacts by 40% at $R_0 = 2.46$ or by 55% at $R_0 = 3.10$. |
| SC4 | Partial lockdown of the country - II | Colleges are closed – reduction in contacts by 75%; reduction in contacts in the workplace by 50%; reduction in contacts of the elderly (older than 65) by 60% at $R_0 = 2.46$ or by 65% at $R_0 = 3.10$; social distancing of the unemployed population and in public places – reduction in contacts by 40% at $R_0 = 2.46$ or by 55% at $R_0 = 3.10$. |
| SC5 | Partial lockdown of the country - III | Reduction in contacts in the workplace by 50%; reduction in contacts of the elderly (older than 65) by 60% at $R_0 = 2.46$ or by 65% at $R_0 = 3.10$; social distancing of the unemployed population and in public places – reduction in contacts by 40% at $R_0 = 2.46$ or by 55% at $R_0 = 3.10$. |

The timing of the simulation of anti-epidemic measures, i.e. the reduction of individual contacts corresponds to the actual date when the implementation of measures in the real epidemic in Serbia began (March 15, 2020). Considering that it is not realistic to expect the desired level of reduction of physical contacts to be achieved in one day, in each scenario, a period of gradual introduction of measures was simulated (lag period of 7 days). Since each individual scenario was simulated at R_0_ of 2.46 and 3.10, different contact reduction rates were applied accordingly.

Additional four scenarios of control of COVID-19 were simulated based on vaccination policy. We assumed that vaccine efficacy was 50%, 68%, 80%, and 85%. The initial conditions assumed that all other anti-epidemic measures are excluded from the model and replaced with mass vaccination. Indicators of epidemic dynamics were monitored, such as: CI, hospitalized patients, patient in intensive care units and deaths.

### 2.5. Model sensitivity analysis and calibration

Considering the world experience with detection of COVID-19 cases, as well as the unreliable data on COVID-19 infections which are currently available worldwide, model calibration is very challenging, and can result in obtaining inaccurate values for the parameters [40]. This is especially due to the facts that a significant percentage of the infected individuals do not exhibit any symptoms. The other issue is small percentage of tested population [40].

As part of the national seroepidemiological study, 1,006 subjects were tested in Serbia from May 11^th^ to June 25^th^, 2020, for the purpose of estimating the extent of COVID-19 infection. According to the published data, seroconversion was confirmed in 6,4% of the subjects. On the other hand, a total of 13,372 cases of the infection were reported, which means that those who were infected constitute around 0.19% of the overall population. However, it is our opinion that the data on reported deaths caused by COVID-19 infection is more reliable for use in model calibration, e.g. infection fatality rate. Alex et al. reached a similar conclusion when simulating COVID-19 by using the SEIRD model with heterogeneous diffusion [40]. When we compared the data recorded during the beginning of the epidemic in Serbia with the results obtained during the simulation, such as the initial doubling time, the two data series matched well. However, later, the obtained results did not match well the officially registered data on the number of infected, especially after the beginning of the implementation of measures in Serbia. We attribute these differences to the methodology by which official authorities register cases of infection, and collect the data.

The parameter that determines the number of deaths is the IFR. It is the number of persons who die of the COVID-19 among all infected individuals regardless of whether the infected show symptoms of the disease or not. As with many diseases, IFR is not always equivalent to the number of reported deaths caused by COVID-19. This is because a significant number of deaths, although caused by COVID-19, will not be recognized as deaths caused by COVID-19 [41]. Also, there are many asymptomatic cases of infection which are never detected [42, 43, 44, 45].

However, according to new findings, the overall estimate of the proportion of people who become infected with SARS-CoV-2 and remain asymptomatic throughout infection was 20% (95% confidence interval) with a prediction interval of 3%–67% in 79 studies that addressed this question [46, 47]. Michael A. Johansson et al. reported that 30% of individuals with infection never develop symptoms and are 75% as infectious as those who do develop symptoms, and concluded that persons with infection who never develop symptoms may account for approximately 24% of all transmission [39].

Due the fact that there is a lag in time between when people are infected and when they die, patients who die on any given day were infected much earlier, and thus the denominator of the mortality rate should be the total number of patients infected at the same time as those who died [41]. David et al. estimated mortality rate by dividing of deaths on a given date by the total number of persons confirmed as COVID 19 cases 14 days before [29, 41]. It is based on the assumption that maximum incubation period is14 days [34]. However, if we take into account that the number of registered cases of COVID-19 infection is usually significantly lower than the actual number, assuming that the data on deaths are accurate, the real IFR value is significantly lower than the calculated value [40]. If we apply this to the situation in Serbia, the daily value of IFR on July 10^th^, 2020, when the largest number of deaths was registered in one day, was 9.33%, considering that 18 deaths were registered on July 10^th^ and 14 days earlier 193 confirmed cases of COVID-19 infection. The raw values of IFR for the period between March 6^th^ and August 10^th^ were as follows: median of IFR = 2.11%, and average value of IFR = 7.15% bounded in interval 4.17%-10.13%. When we compared these values with those published by the WHO, CDC and other authors [43, 44, 48] we concluded that they differ significantly. Considering these findings, the IFR values adopted in the model (for various population groups and genders) were primarily taken from the literature, with a remark that the selection of IFR values was based on preliminary comparison of the overall Serbian IFR with similar IFRs found in the literature, taking into account the registered deaths and most probable number of infected individuals [33]. To make this possible, the first step was to correct the local raw IFR value mentioned above. Based on real data we first calculated the population at risk of dying from COVID-19 infection for each individual day since the outbreak, ending on August 10^th^, 2020. The number at risk on a given day should correspond to the number of deaths from COVID-19 infection, considering the lag period from infection to death. For this calculation, we used the data on the number of deaths *D*_*t*_ in Serbia registered on daily bases [2]. We hypothesized that the distribution of time periods *t*_*n*_ from the moment of COVID19 infection to death follows the lognormal distribution defined by the parameters *m* = 26.8 and σ = 12.4 days [48, 49].

Based on the formula: 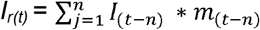 by reverse, we calculated the population at risk of dying from COVID-19 infection for each individual day, where *m*_*t*_ is the probability that the time between infection and death is *t* days and follows the lognormal distribution (*m* = 26.8, σ = 12.4) [48, 49]. After that, the daily IFR values were calculated according to the formula IFR_(t)_ = D_t_ /I_*r(t)*_ [50]. Based on IFR values calculated in this way, we made descriptive statistics and obtained the mean value of IFR = 0.70%, bounded in the interval 0.46-0.94% and a median of 0.19%. It is important to note here that this value corresponds with the COVID-19 IFR values found in Eastern European countries and Spain [50, 33]. Taking these findings into account, we decided to take the IFR values specific to certain population strata recorded in Spain as the most appropriate for our case [33]. The adopted IFR values are listed in Table 3.

In this section, we used sensitivity analysis to estimate the amount of change on outcomes when varying the input values used in the model. Sensitivity analysis is one of the methods most frequently used for the evaluation of disease spread models [51, 52]. A sensitivity analysis is carried out to characterise the impact of uncertainty of input parameters on model outputs. Sensitivity analysis consists of assessing the impact that changes in input parameters have on model outcomes. We evaluate two aspects of the model: the global behavior of the model when perturbing a group of key parameters together, and the impact of changes when perturbing the key parameters used in the model individually.

The model sensitivity analysis was conducted by changing the most sensitive model parameters: *R_0_, β, f-1, r, b, m, γ, ε, η, ω*, 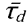. The values of these parameters were increased by 5%, 10% and 25% relative to the base scenario and changes in output indicators were observed.

Finaly, validation of the model was performed by comparing the historical data of the real epidemic in Serbia with the data obtained with the SEAIHRDS model. For validation purposes, the current epidemic of COVID-19 was simulated, along with actual anti-epidemic measures. Taking into account the risks described above related to the accuracy of historical data from the COVID-19 epidemic in Serbia and significantly higher confidence in the accuracy of data related to the number of deaths from COVIDA-19 compared to data on the number of infected, for the purpose of model validation, only data on the cumulative number of deaths were used. The reason for this assumption is that there is still uncertainty about the proportion of the infected population that is not reported due to a mild form of the disease or the patients are asymptomatic.

In statistical analysis the coefficient of determination, R^2^, was used to check goodness of fit of SEAIHRD model with COVID-19 data recorded during the real epidemic. The regression coefficient compares predicted values (y) against actual data (x). To address model uncertainties, bias, mean absolute error (MAE), mean square error (MSE), the root mean square error (RMSE), maximum deviation (MaxDev) and normalized root mean square error (NRMSE) were also estimated (*Supplementary material*).

## 3. Results

### 3.1. Predicting the number of sick, hospitalized patients and deaths caused by COVID-19 in the absence of any intervention measures

After the simulation, the model predicted that with R_o_ = 2.46, and without the implementation of any anti-epidemic measures, the initial doubling time of the infection could be five days. The epidemic wave could peak 219 days after the outbreak, and it could yield 99,819 infected individuals in a day. Afterwards, the infection rate could decline for 215 days, eventually reaching the daily incidence of 492 newly infected, after which the next epidemic wave could start. The second wave could peak 706 days after the onset of epidemic and yielding 25,232 infected individuals in a day. The third epidemic wave could peak 429 days later, with 13,709 infected individuals in a day. The true cumulative incidence in the first year of the epidemic could be 6,229,144 infected people with SARS-Cov-2 virus, while the apparent cumulative incidence could be 4,360,401 infected. A total of 20,894 patients could die due to COVID-19 consequences.

With R_o_ = 3.1, the following results were obtained: the initial doubling time of the infection was five days, true cumulative incidence 7,133,221, apparent cumulative incidence 4,993,254, and the total deaths of 23,951. Fig. 1, panels a) and b) show daily variations in the number of susceptible, latently infected, infected and recovered patients, at basic reproduction numbers of R_o_=2.46 and R_o_=3, respectively. Panels c) and d) of the same figure show daily fluctuations in susceptible, recovered and net reproduction rates R_n_ for R_o_=2.46 and R_o_=3, respectively. Panels e) and f) of Fig. 1 show daily variations in R_n,_ true and apparent disease incidences at basic reproduction numbers of R_o_=2.46 and R_o_=3. The shaded area corresponds to the period when the daily number of new COVID-19 infected individuals increasing, and therefore all of the following hold: R_n_>1, proportion susceptible >1/R_o_ and the proportion of population that is recovered (immune) is below the herd immunity threshold. Fig. 2 panels a) and b) show a prediction of necessary hospital capacities. Panels c) and d) of Fig. 2 show predicted numbers of sick and dead due to COVID-19 at R_o_=2.46 and R_o_=3 and age structure of hospitalized patients and deaths.

**Fig. 1.**
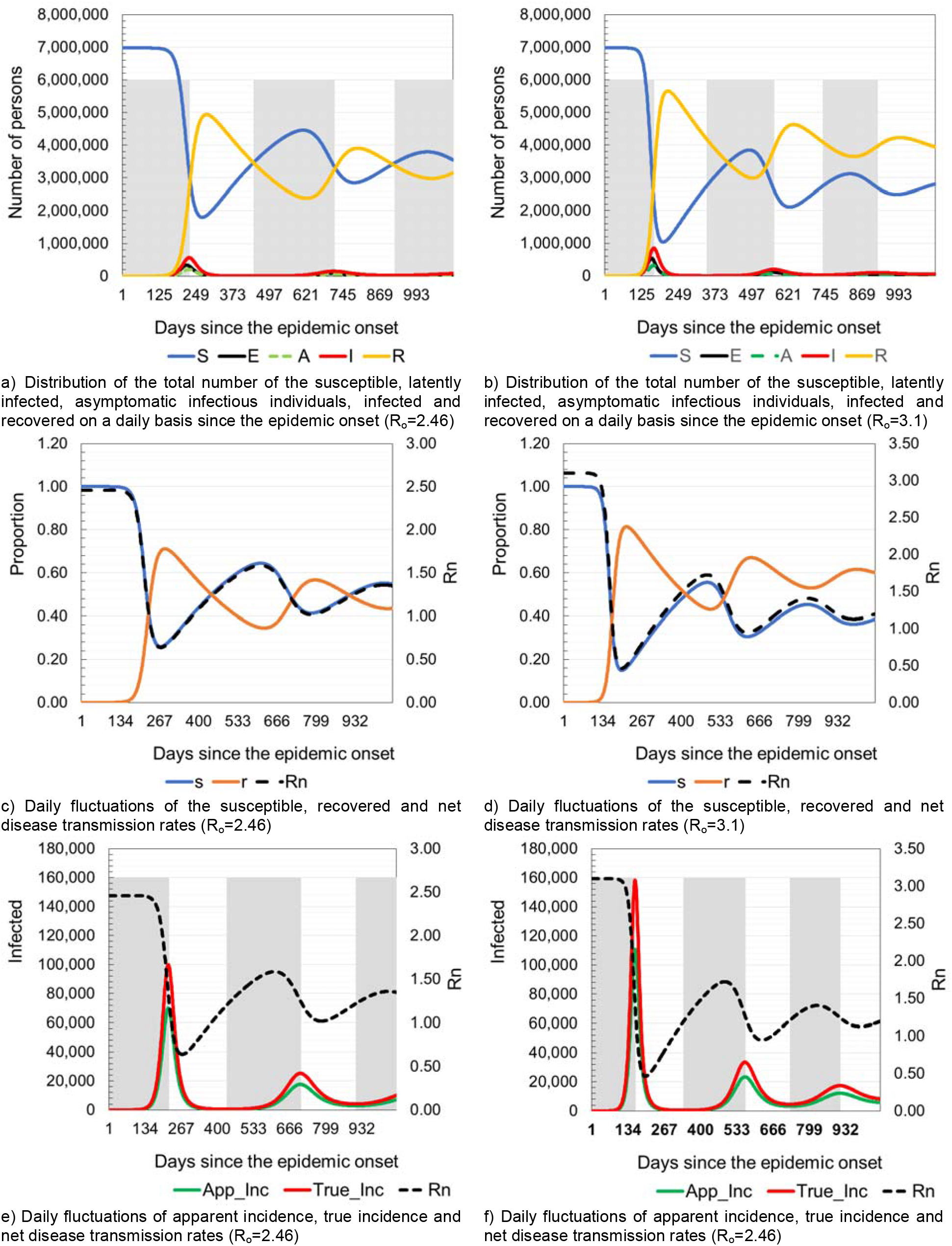
Model prediction of latently infected, asymptomatic infectious individuals, infected, recovered and daily fluctuations of R_n_.

**Fig. 2.**
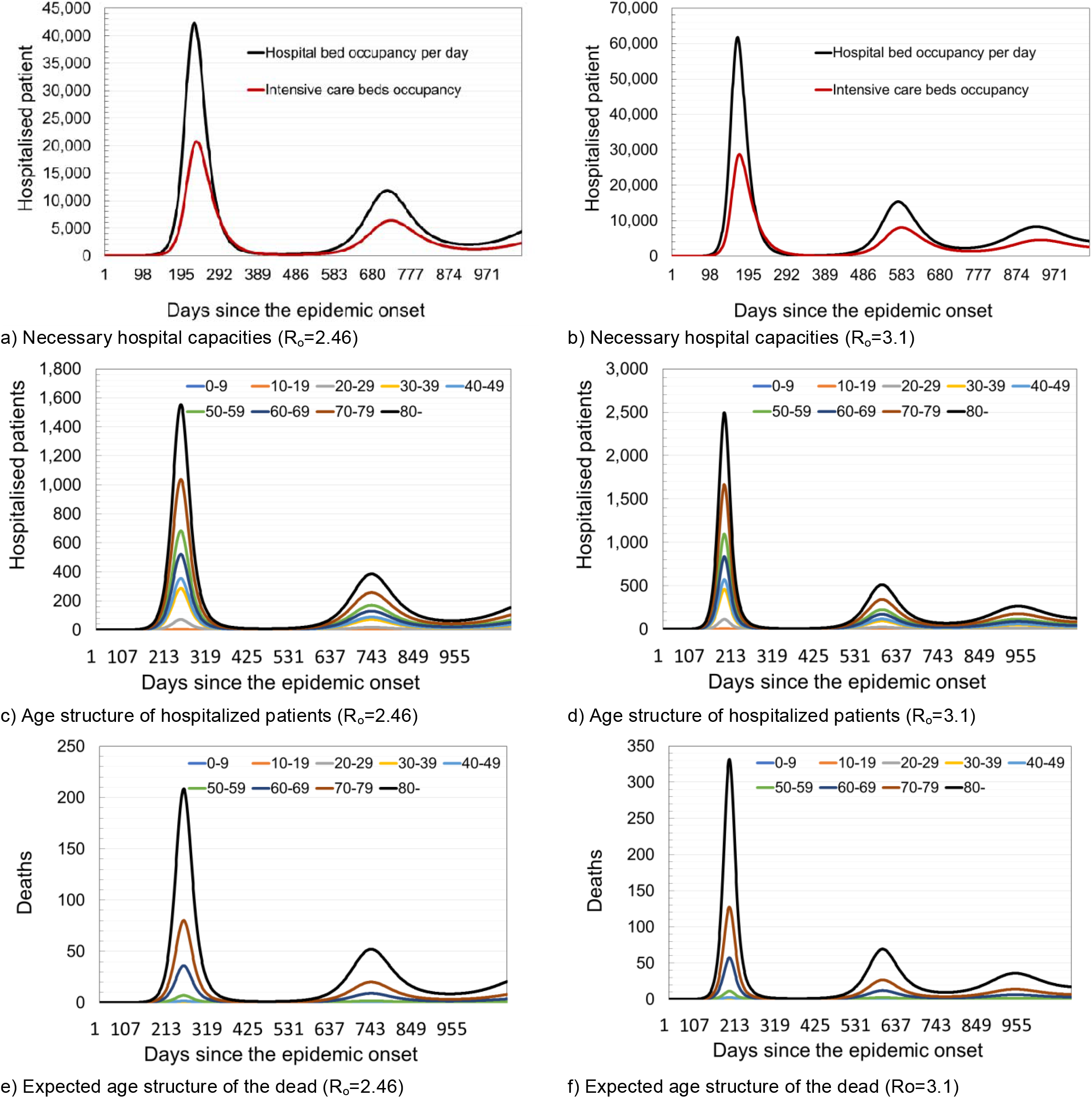
Model prediction of required hospital capacities under the assumption of different intervention measures.

### 3.2. Predicting the number of sick, hospitalized patients and deaths caused by COVID-19 in the conditions of application of restrictive anti-epidemic measures

When the spread of COVID-19 epidemic through totally susceptible population in the Republic of Serbia is simulated, under an assumption of only incidental movement among the population, basic reproduction number of R_o_ = 2.46, and with lock-down of entire country, a significant slowdown of the epidemic was observed. Initial infection doubling time was 6 days. The peak of the epidemic wave could occur 702 days after the epidemic onset, when there could be 2,848 infected in one day. In the first year of the epidemic 308,581 people could be infected and 1,031 people could die.

When the basic reproductive number was increased to R_0_ = 3.1, and for certain segments of the population the contact reduction factor *ρ*_*i*_ increased compared to the values from the scenario with R_0_ = 2.46 (reduction of contacts in public places and contacts of persons over 65 years by 55% and 65%, respectively), the results changed significantly. The model predicted that with R_0_ = 3.10, the initial infection doubling time could be 5 days, the peak of epidemic wave could occur after 246 days and it yield 8,110 infected people in one day. In the first year of the epidemic 2,219,251 people could be infected and 7,194 could die.

Table 5 provides the overview of epidemic indicators obtained from the simulations of all five scenarios with R_o_ = 2.46 and R_o_ = 3.1. Figures 3. and 4 provide a comparative overview of results of all five simulated scenarios. Panels a) and b) of Fig. 3 show the values of cumulative incidences on a daily basis for R_o_=2.46 and R_o_=3.1, respectively. Panels b) and c) of Fig. 3 show the expected total number of hospitalized patients and patients in intensive care units (ICU) for R_o_=2.46 and R_o_=3.1, respectively. Fig. 4 provides overview of required hospital capacities e.g. hospital bed occupancy and the occupancy of beds in ICU on daily bases for R_o_=2.46 and R_o_=3.1, respectively. The results show that after applying various measures to slow down the circulation of SARS-Cov-2, the number of newly infected people, hospitalized patients and the occupancy of hospital capacities are the lowest in the situations where rigorous anti-epidemic measures are applied to all population strata (Scenario 2 in Table 5 and in Fig. 3 and Fig. 4). Openings of pre-school and elementary school’s facilities leads to a visible jump in the number of infected and hospitalized in all strata. This finding clearly shows that children, although least susceptible to developing more severe clinical pictures, are important when transmitting SARS-Cov-2 (Scenario 3 in Table 5 and Fig. 3). Opening of the high schools and colleges also leads to a visible increase in the number of newly infected and hospitalized patients, including an increase in the number of deaths (Scenario 5 in Table 5 and Fig. 3). Without the application of any intervention measures, the greatest burden on the health system could be expected 228 days from the beginning of the epidemic at R_o_=2.46 or 168 days at R_o_=3.1.

**Table 5.**
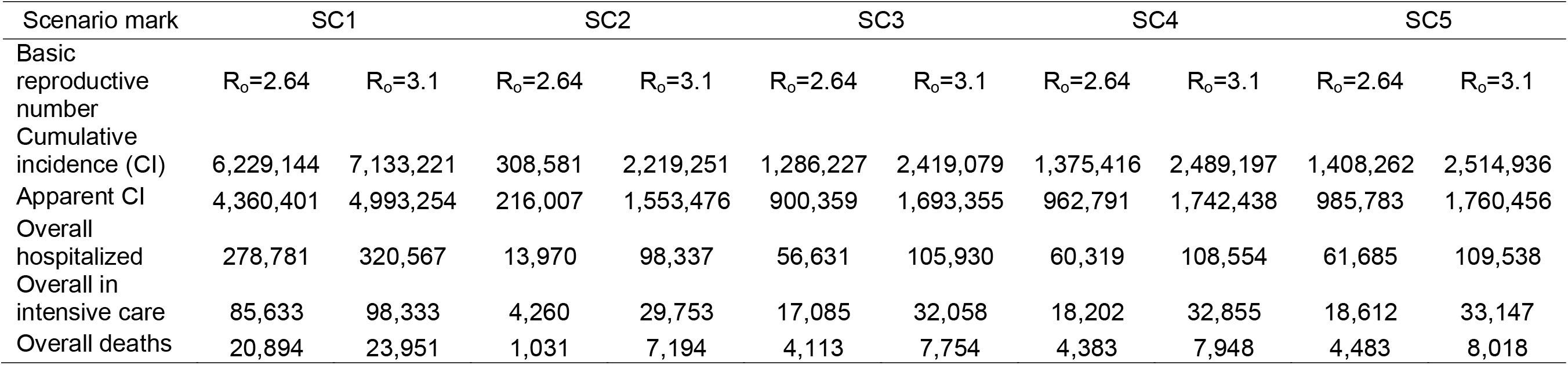
Results of different simulated scenarios (Ro = 2.46 and Ro = 3.1). The data refers to the period of 365 days from epidemic onset.

**Fig. 3.**
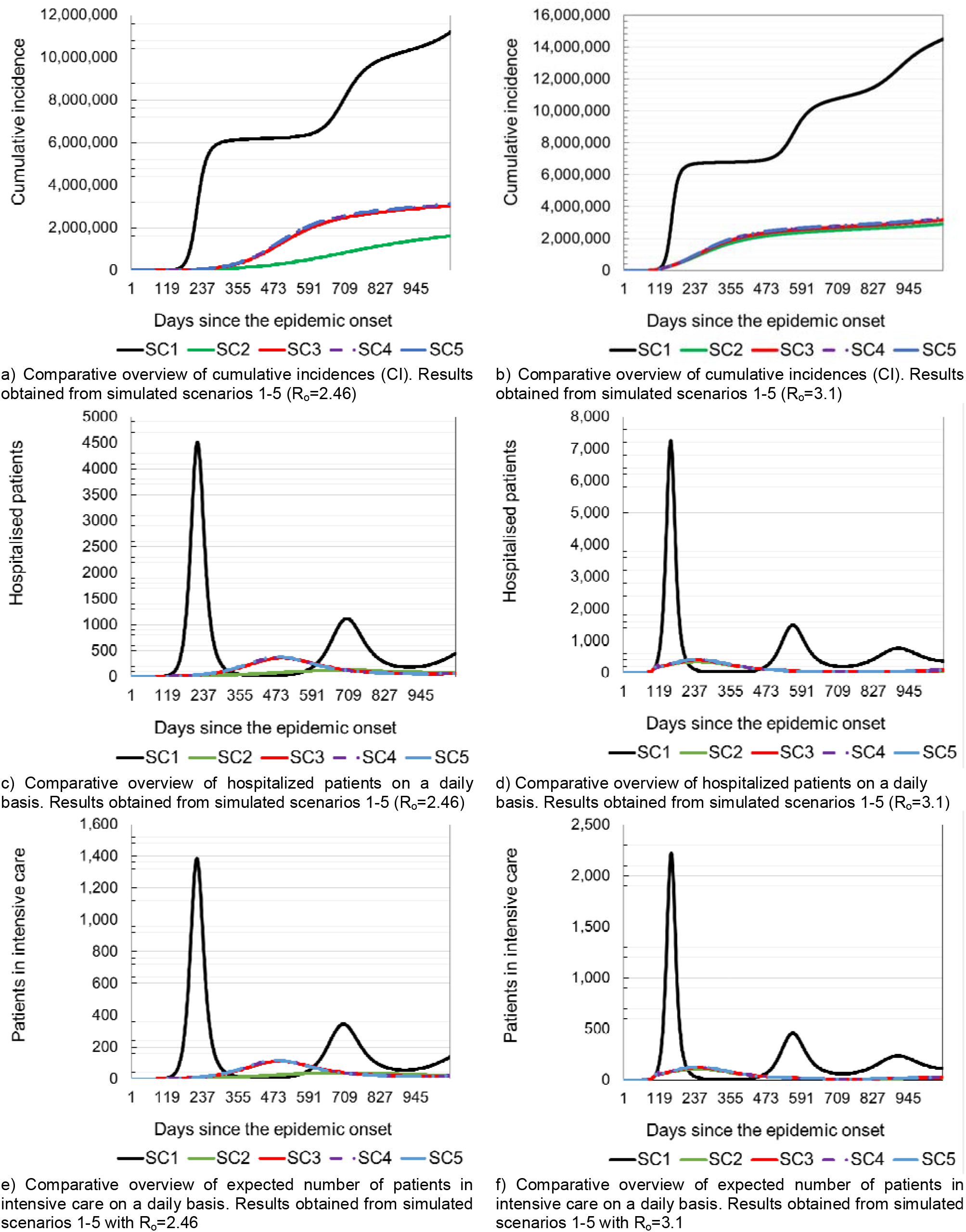
Model prediction of expected number of hospitalized patient and patient in intensive care.

**Fig. 4.**
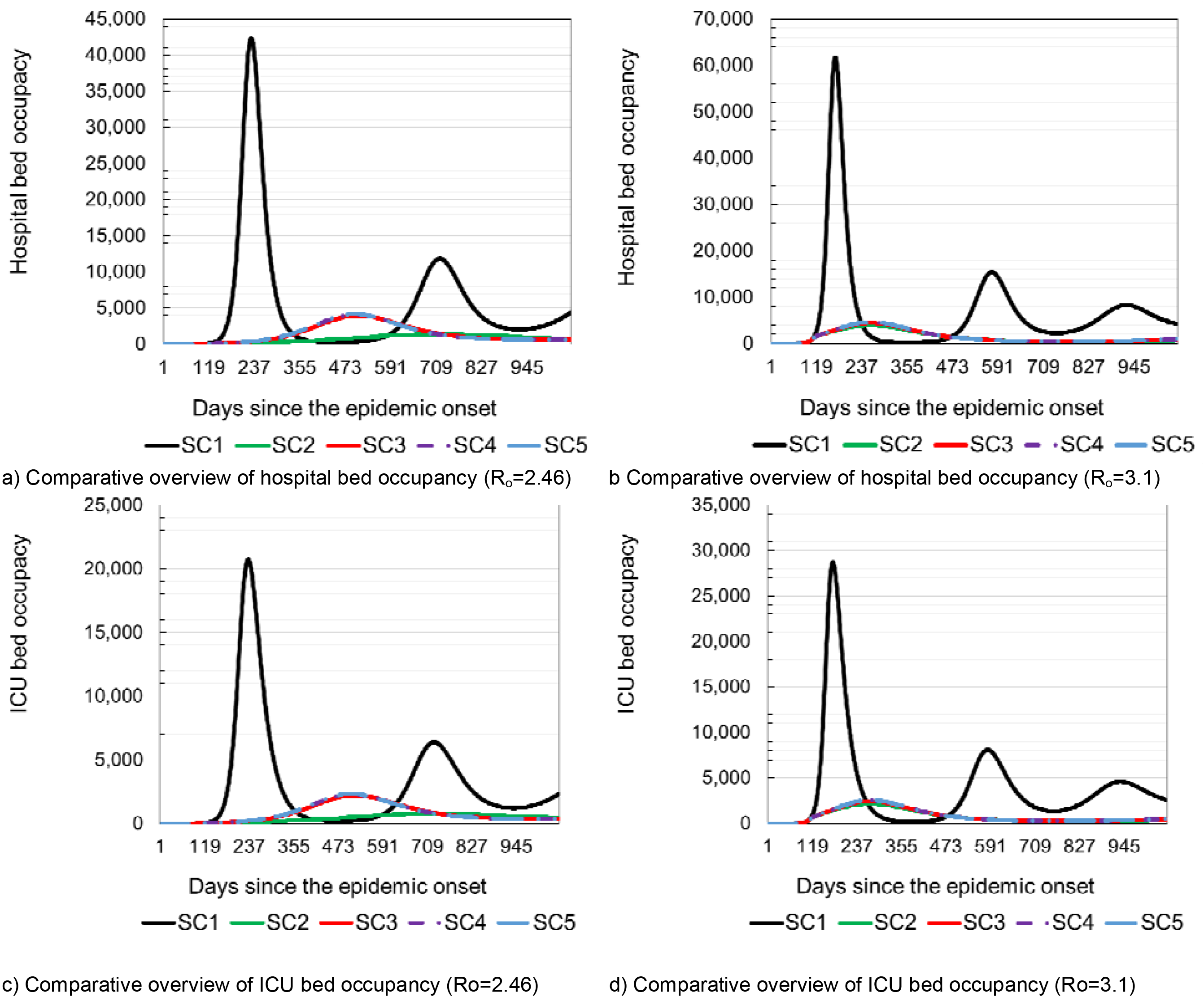
Model prediction of required hospital capacities needed to treat patients with COVID-19.

Depending on the R_o_ value used in simulation, it could be necessary to provide 42,351 (61,739) hospital beds and an additional 20,173 (28,740) in intensive care units. In the case of Scenario 2, there is a significant slowdown in the epidemic. According to the predictions obtained by the simulation of Scenario 2, after 713 (259) days at the moment of maximum occupancy of hospital capacities, it might be necessary to provide 1,387 (3,940) beds in COVID-19 hospitals and 784 (2,162) beds in intensive care units (Fig. 4).

### 3.3. COVID-19 simulation and disease control by implementing a hypothetical vaccine

Assuming that the disease is spreading at the basic reproduction number of R_o_=2.46, the herd immunity threshold (when the disease can be expected to slow down and the chain of infection is expected to be broken) would be 59.35%, while at R_o_=3.1, it would be 67.74%. Depending on the efficacy of the potential vaccine, the required vaccination coverage should be 87% (*v*_*e*_ =68%), 74.19% (*v*_*e*_ =80%), 69.82% (*v*_*e*_=85%), 65.94% (*v*_*e*_=90%) and 95.41% (*v*_*e*_=71%), 84.68% (*v*_*e*_=80%), 79.70% (*v*_*e*_=85%), 75.27% (*v*_*e*_=90%) for R_o_=2.46 and R_o_=3.1, respectively. The different ways of including vaccination in the SEAIHRD model are detailed in the supplementary material. Fig. 5 shows different scenarios of COVID-19 control strategies based solely on vaccination.

**Fig. 5.**
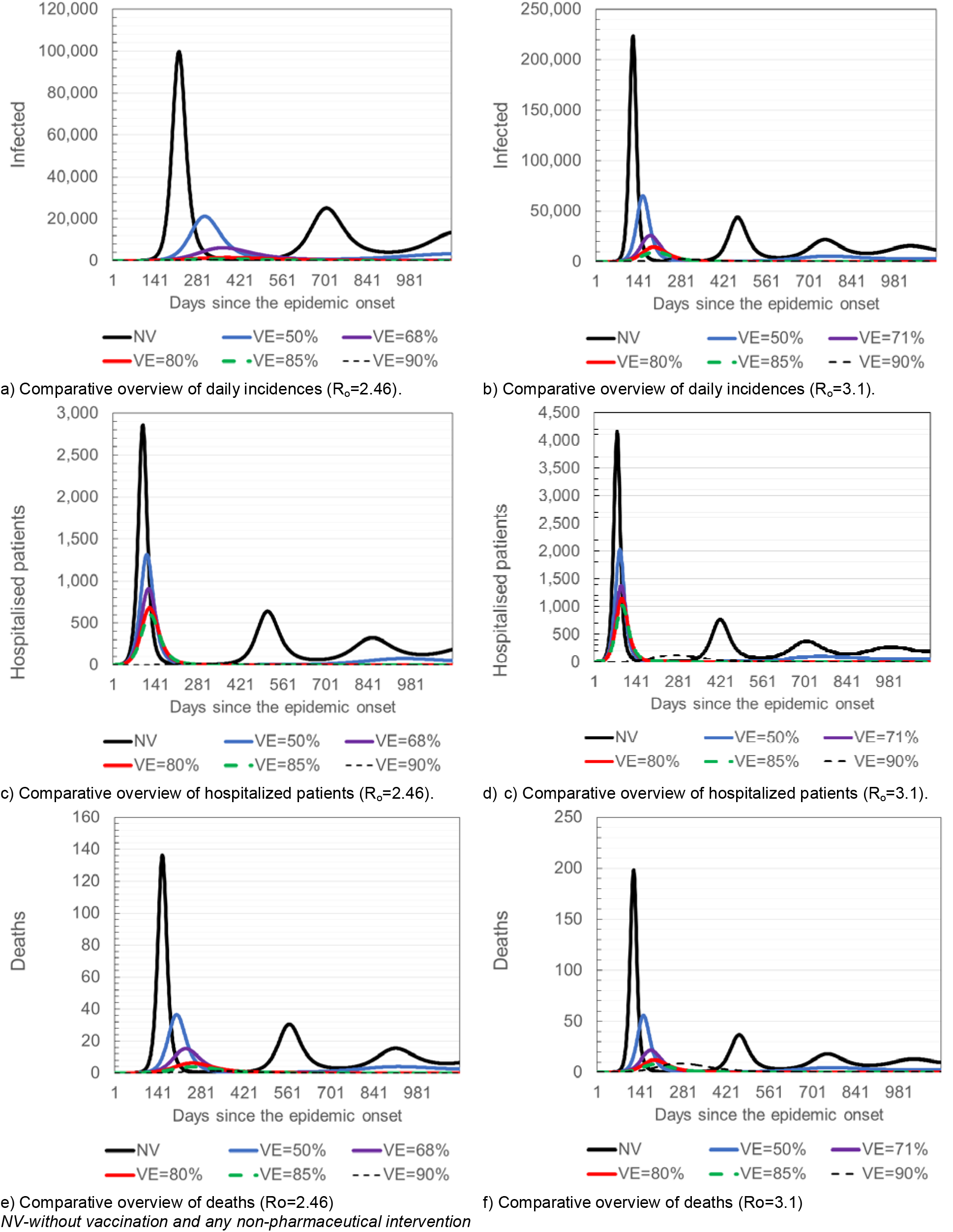
Results of simulated COVID-19 control based solely on vaccination, scenarios 6-9 (R_o_=2.46 and R_o_=3.1).

### 3.4. Results of the model sensitivity analysis

The results of the sensitivity analysis are presented in tables 6 and 7. The tables show increased values of input parameters and the percentage of the parameter increase relative to the basic scenario, as well as the values of output results obtained after the simulation of the changed scenario.

**Table 6.**
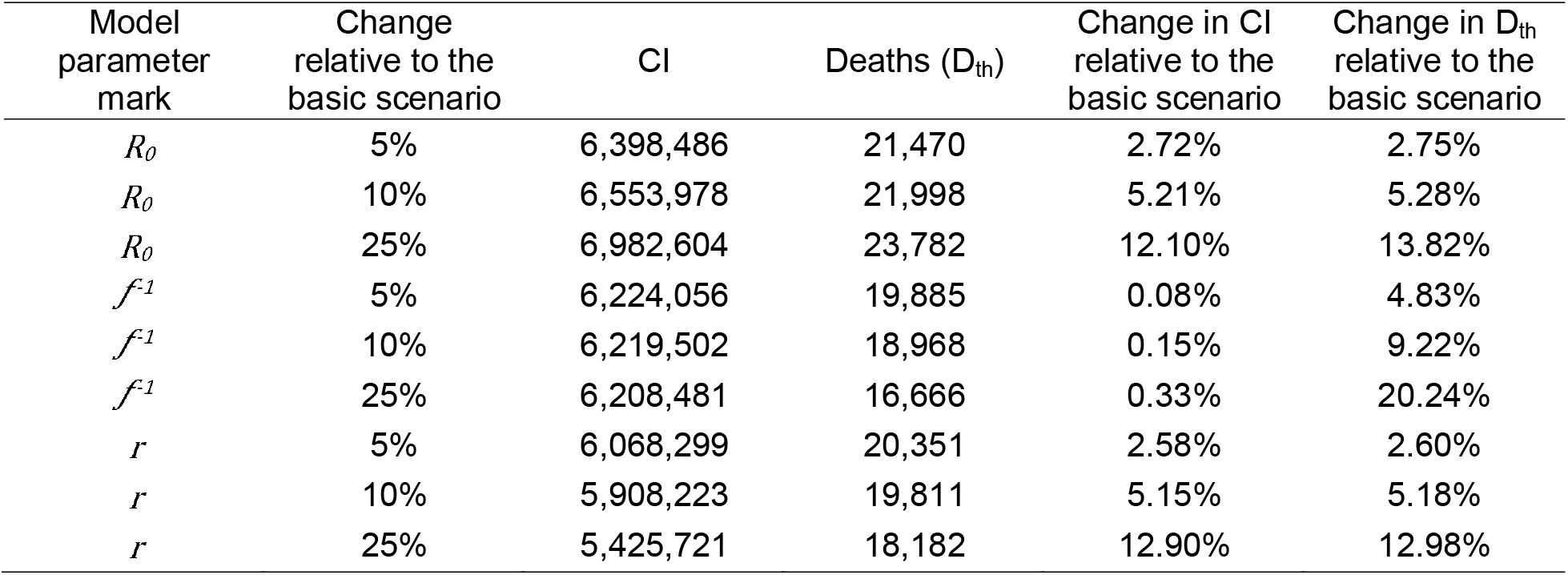
Results of the model sensitivity analysis of individual parameters used in the model

**Table 7.**
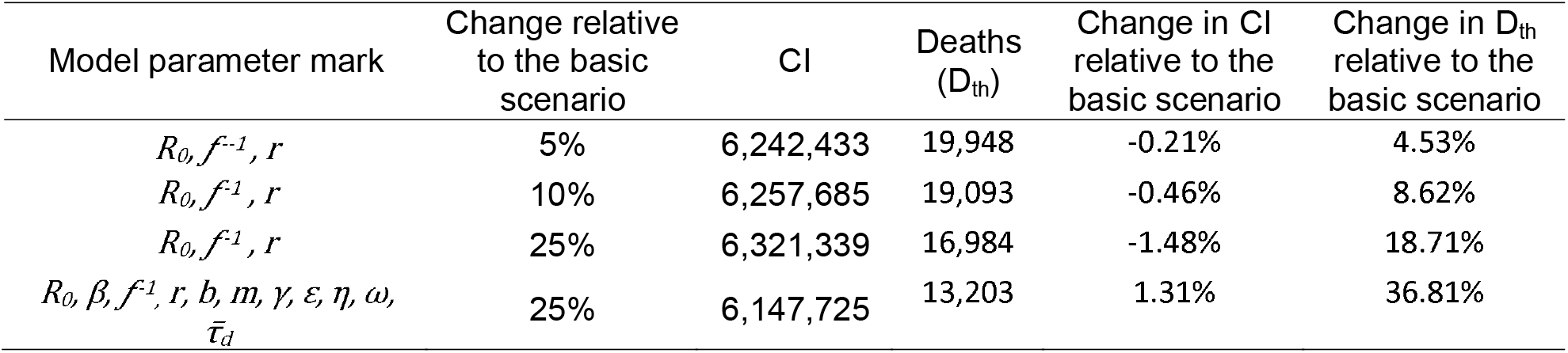
Results of the model sensitivity analysis of group of parameters using a perturbation up to 25%

### 3.5. Results of the model validation

The model was validated on the most recent historical data (from January 1, 2021 to February 1, 2021). As shown in Table 8. all measures of the prediction quality of deceased due to COVID-19 are low. The average difference between the actual number of people who died of COVID-19 and predicted one is only 2.05% and the maximum deviation between the predicted and actual number will not exceed 4.82% with a probability of 95%.

**Table 8.**
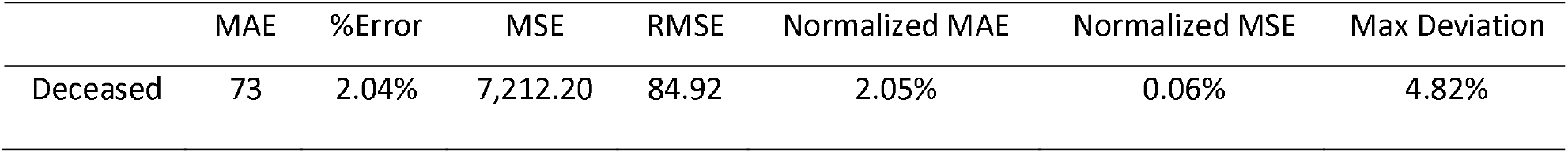
Measures of the prediction quality

Based on Fig. 6 and Table 9, we can conclude that the SEAIHRD model fits historical data quite well. Pearson’s r and coefficient of determination (R^2^) have shown strong the linear relationship between real deceased and the number of deaths predicted by SEAIHRD model.

**Fig. 6.**
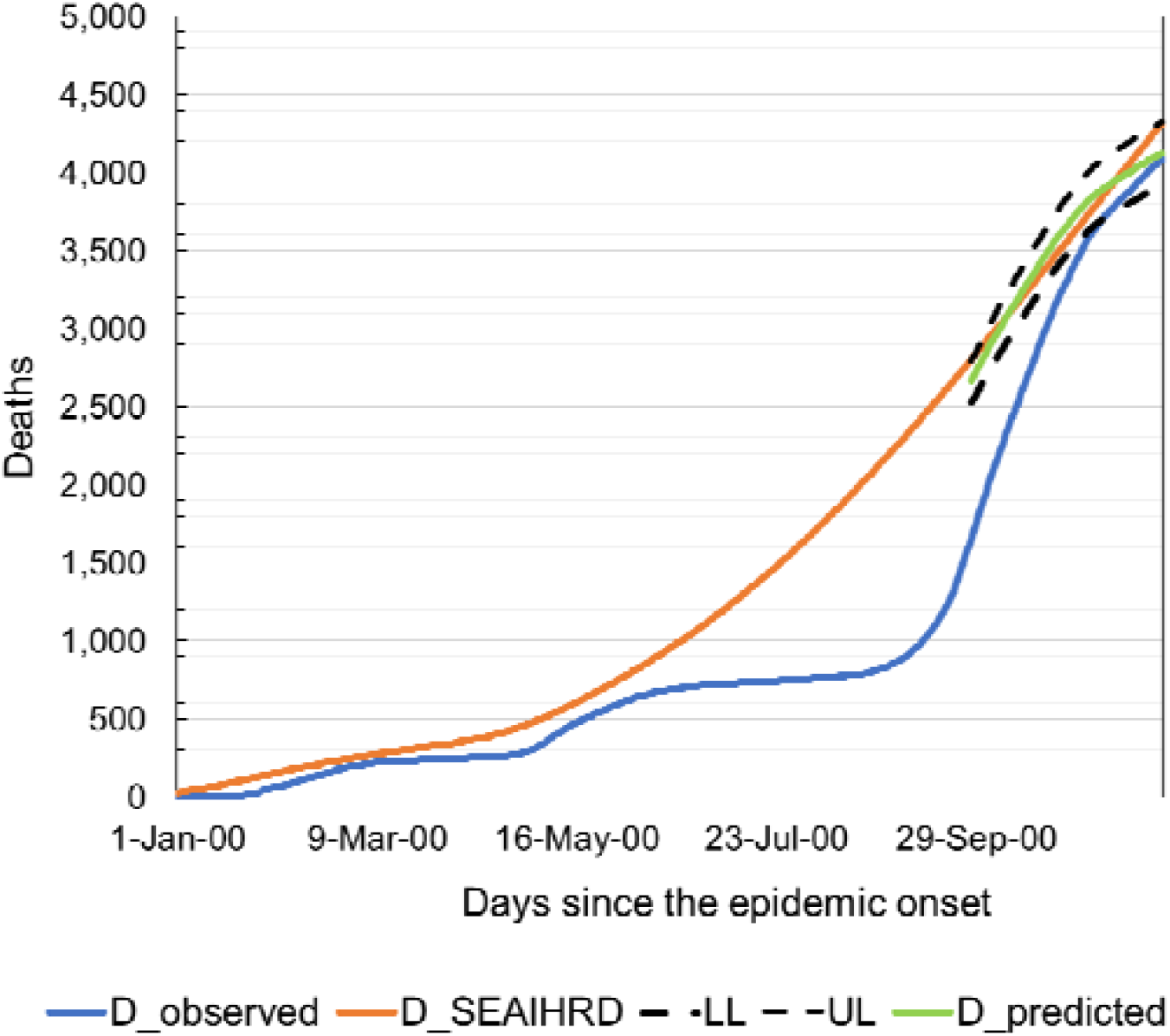
The observed number of deceased individuals (blue), number of deaths modeled with SEAIHRD model (yellow) and predicted number of deaths (green).

**Table 9.**
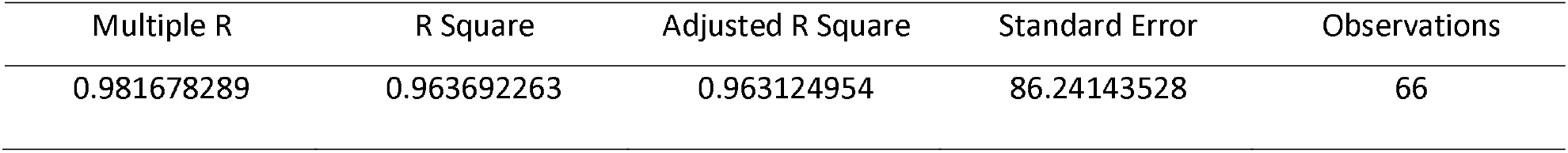
Regression statistics

| Multiple R | R Square | Adjusted R Square | Standard Error | Observations |
| --- | --- | --- | --- | --- |
| 0.981678289 | 0.963692263 | 0.963124954 | 86.24143528 | 66 |

More information on validation results are detailed in the supplementary material.

## 4. Conclusions and discussion

For the needs of this research, we augmented the classic deterministic model by adding the compartments of vaccinated, asymptomatic, hospitalized and latently infected subjects. By adding birth and death rates, we enabled daily fluctuations of the overall simulated population, which brought us closer to the real conditions in which the disease is transmitted. When we assumed that the recovered lose immunity over time, we obtained dynamic oscillations of epidemic waves through susceptible population.

The input values for the parameters used to simulate the COVID-19 were obtained either from literature review or were calculated on the basis of data have taken from the literature and other official sources. Whereas some of these inputs are well documented; other input values are either not so well documented or based on potential subjective opinions (i.e., expert opinion, historical data from epidemic etc.). In any case, well documented or not input values have potential to impact results and, therefore, should be carefully evaluated. The results of conducted sensitivity analysis show that the sources of uncertainty are different for each output considered and it is necessary to consider multiple output variables for a proper assessment of the model. The most influential parameter is *r* than R_0_ and *f*^*--1*^.

The model was tested on Serbian COVID-19 statistic data and obtained validation results allow us to conclude that the proposed model has good prediction ability and performance. However, although we obtained satisfactory results during the validation of the model, worth noticing also that some of the model parameters were estimated based on available data that might be less precise due to the difficulty of being measured. That could be the reason why the values of some parameters e.g. recovery rate, contact rate, daily infection rate, that are estimated in hospitals may differ from those acquired by this study. For that reasons SEAIHRD model can be used for the long-term rough predictions of the epidemic. Obtained long-term predictions reflect the general dynamic of the outbreak and are especially useful for the healthcare system workers and governmental officials.

When we simulated different disease control scenarios of the COVID-19 epidemic based on non-pharmaceutical intervention measures, scenario number 2 proved to be the most effective approach to the disease control, because it implemented the most comprehensive anti-epidemic measures (entire country lock down). However, the basic problem of this approach is the feasibility and practicability to maintain the measures in the long term.

The model predicted that students, children and younger school-age generations have an important role in transmitting COVID-19, especially if they come into contact with a more vulnerable population. The model showed that, in the case of returning school children of all ages to schools, an increase of 10.48% in the estimated deaths and 12.16% of the number of infected is possible, when compared to the conditions before opening of the schools (Scenarios 2 and 4). However, most dead and seriously diseased people are found in the older population. This is particularly important when planning intervention measures, especially when deciding on which restrictions to be lifted and how (opening schools, students’ return to faculties etc.). The model predicted that COVID-19 has a potential to spread rapidly and linger in population. Due to a large number of the infected persons and duration of the disease, there are significant needs for hospital capacities, especially in the conditions when the disease is suppressed by implementing partly relaxed anti-epidemic measures, or in the case of absence of any measures. According to the prediction, without the application of any intervention measures, at the moment of the greatest load, depending of actual R_o_, the health system should provide 42,351 (R_o_=2.46) hospital beds for the care of the patients and an additional 20,733 in intensive care units. On the other hand, in the case of the application of the strictest anti-epidemic measures, the needs decrease to only 1,387 beds in COVID-19 hospitals and additional 784 beds in intensive care units. In the case of continued implementation of current measures, which are significantly less intense than the measures applied at the beginning of the epidemic, it is necessary to provide 3,537 beds in COVID-19 hospitals and 1,945 beds in intensive care units in the entire country. The model also shows that the needs for hospital capacities decline with the ending of the first epidemic wave, since daily incidence decreases and during the second and third waves it never reaches the initial peaks, but these needs still remain substantial. For example, in the case of Scenario 1, at the top of the second epidemic wave, it is necessary to provide 11,845 beds in COVID-19 hospitals and 6,378 in intensive care units, which makes 27.96% and 30,76% of the required capacities of the first wave of the epidemic.

Based on the cyclical patterns of the epidemic waves and duration of simulated epidemics, the model predicted that the disease has a potential to linger in population and that it will most probably have a seasonal pattern. Therefore, potential vaccines can have an enormous potential and significance for COVID-19 control. Depending on the efficacy of future vaccines, the disease can be stopped and curbed almost solely by implementing the measure of vaccination. However, the necessary conditions for these predictions and expectations are the efficacy of potential vaccines and the ability of a health systems to implement vaccination to a satisfactory extent and rapidly, especially with regards to the most sensitive categories of population. Depending on the R_o,_ a vaccine that would have an efficacy ≥ 68-71% could stop the pandemic and break the chain of infection. However, even vaccines with lower efficacy could be useful as they would significantly reduce the number of cases and deaths, especially if used in combination with the other disease control measures. The effects of vaccination depend primarily on: 1. Efficacy of available vaccine(s), 2. Prioritization of the population categories for vaccination, and 3. Overall vaccination coverage of the population, assuming that the vaccine(s) develop solid immunity in vaccinated individuals. With expected basic reproduction number of Ro=2.46 and vaccine efficacy of 68%, an 87%-coverage would be sufficient to stop the virus circulation. The required minimal vaccination coverage should be 87% (*v*_*e*_ =68%), 74.19% (*v*_*e*_ =80%), 69.82% (*v*_*e*_=85%) and 95.41% (*v*_*e*_=71%), 84.68% (*v*_*e*_=80%), 79.70% (*v*_*e*_=85%) for R_o_=2.46 and R_o_=3.1, respectively. The minimum daily vaccination rate should be 0.47% for vaccines with an efficiency of 85%, and 0.59% for vaccines with an efficiency of 68%.

Based on the obtained results, we can conclude that at this point, without the application of specific pharmaceutical products, COVID-19 suppression is highly dependent on the basic reproduction number (R_o_), and that more intensive contacts and relaxed measures can result in a dramatic spread of the virus. The choice of intervention measures depends on the feasibility of their implementation and their efficacy in different social contexts. COVID-19 will likely have to be suppressed in this way for a certain period of time. This will most probably last until sufficient quantities of a reliable and effective vaccine are available, and thereafter until optimal vaccination coverage is achieved.

## Supporting information

Highlights

## Data Availability

The data used to support the findings of this study are included within the article and can be used without restriction.

---

**Fig. 1.**
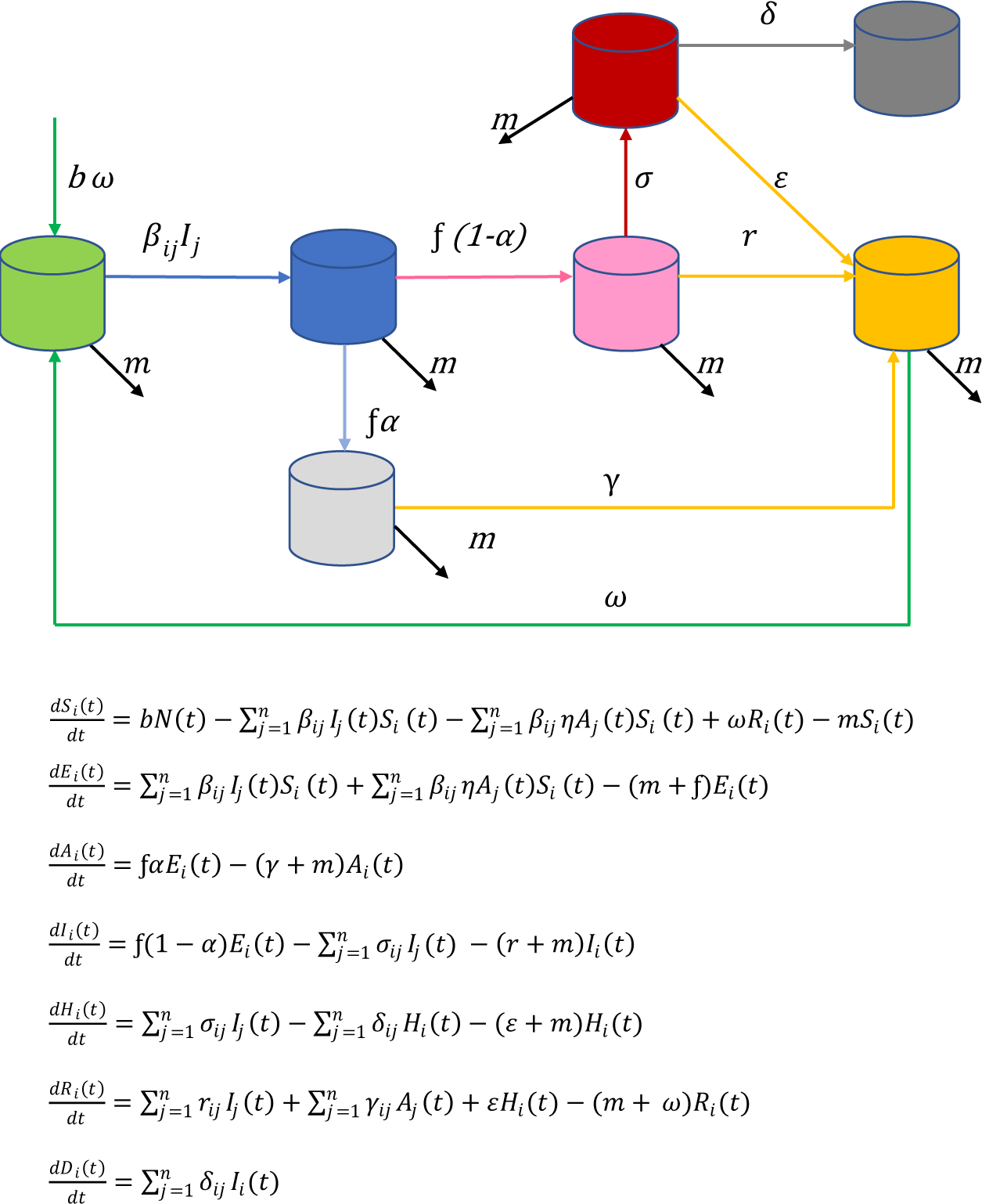
SEAIHRDS model with demography

**Fig. 2.**
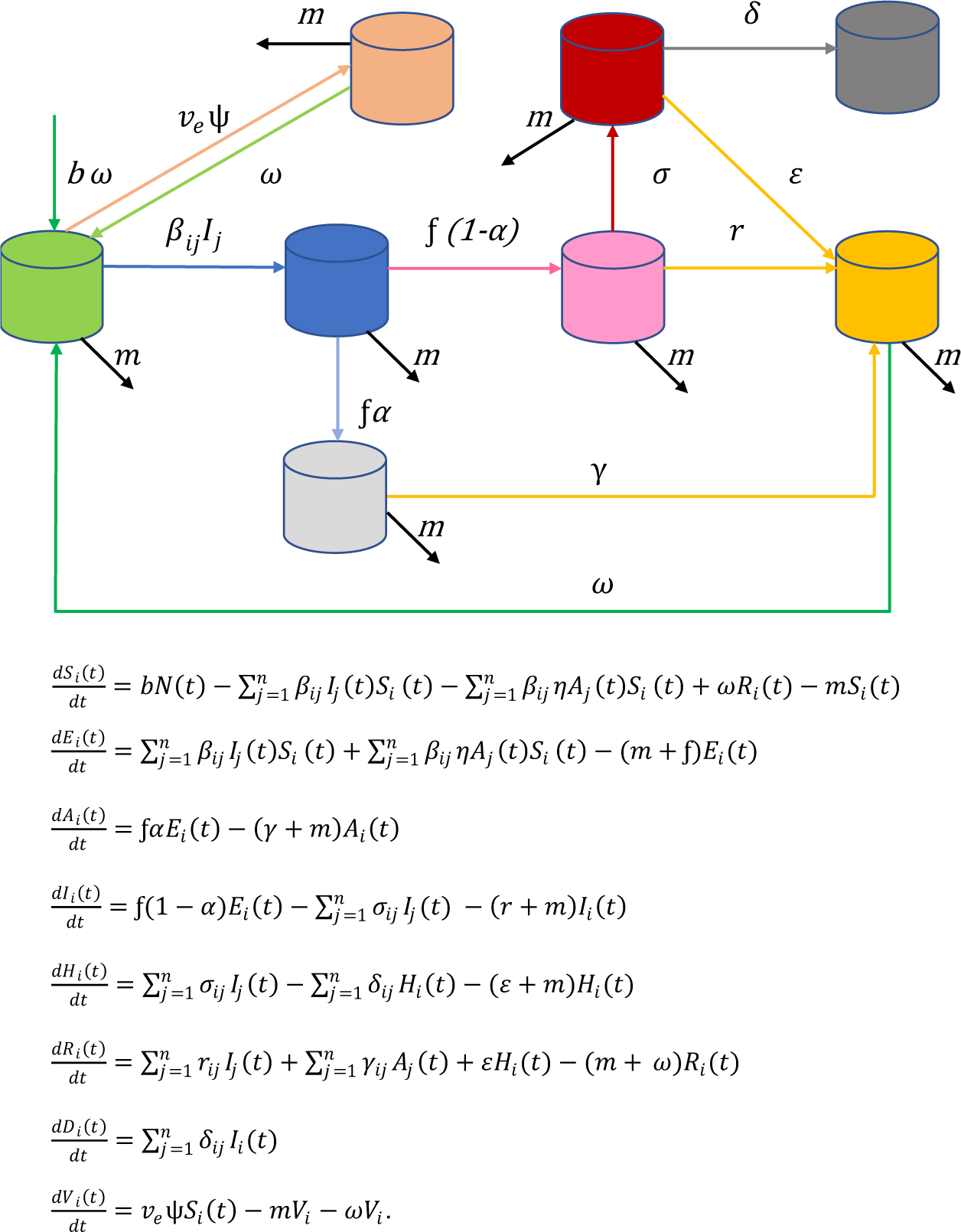
SEAIHRDVS model with demography and vaccination

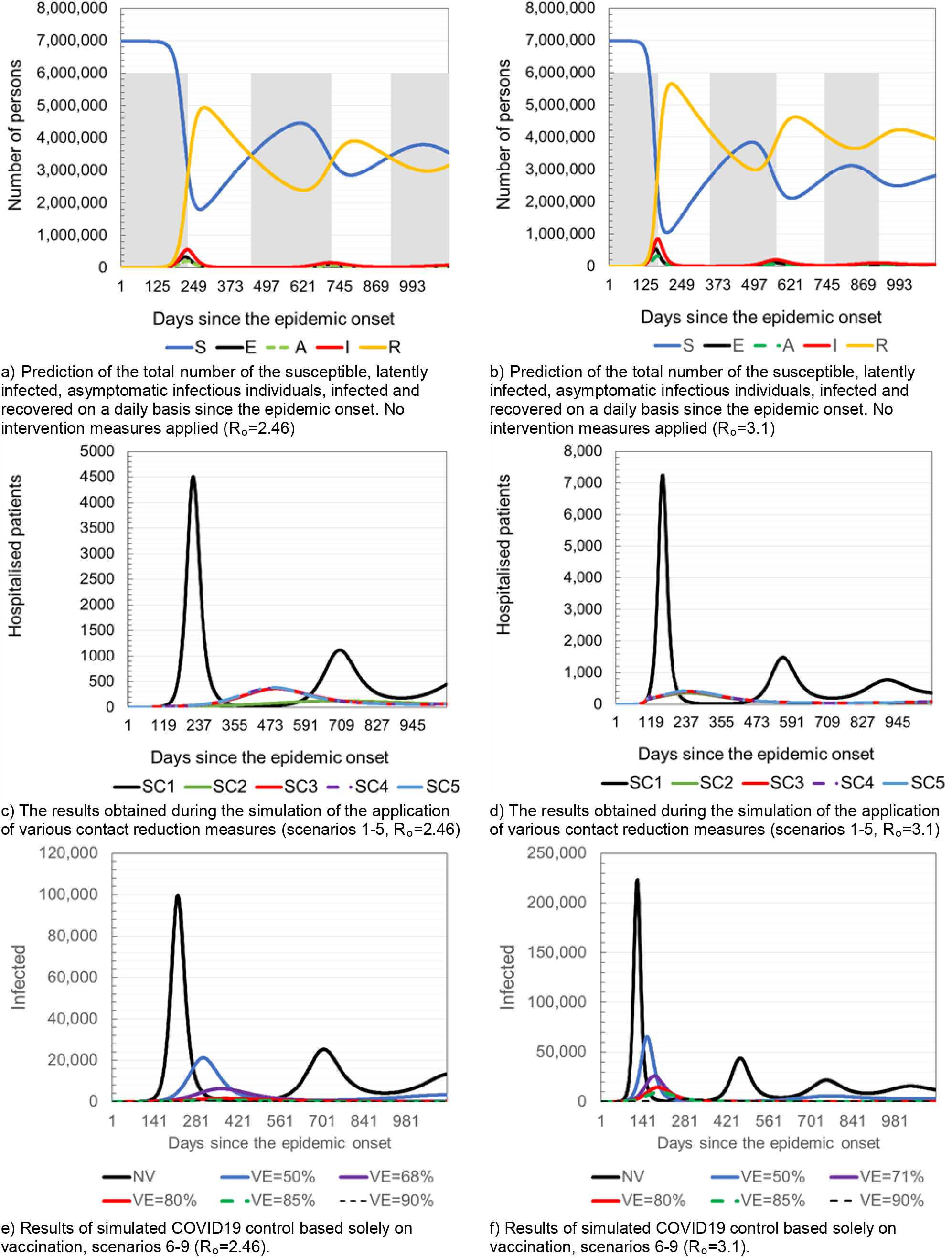

