## Supplementary material for "Simulation and prediction of spread of COVID-19 in The Republic of Serbia by SEIRDS model of disease transmission": Highlights

An augmented multi-compartment SEAIHRDS model was used to analyse different scenarios of COVID-19 pandemic and required hospital capacities.

The model demonstrates that COVID-19 has a potential to spread rapidly and linger in the population and probably will have a seasonal pattern.

Depending on the efficacy of future vaccines, the disease can be stopped and curbed almost solely by implementing the vaccination.

The necessary conditions for successful control of COVID-19 by vaccination in the future are the efficacy of potential vaccines and the ability of a health system to implement vaccination to a satisfactory extent.

Without the application of specific pharmaceutical intervention, COVID-19 control is highly dependent on the basic reproduction number ($R_o$), and the relaxation of control measures can cause a dramatic spread of the virus.
