## Supplementary material for "Simulation and prediction of spread of COVID-19 in The Republic of Serbia by SEIRDS model of disease transmission"

### 1. Description of SEAIHRDS model

Classical SEIRDS model divides the population into compartments, i.e. groups. The population is divided into the following compartments: susceptible to the infection (S), those latently infected with SARSCov-2 (exposed to) (E), the infected individuals who are able to spread the disease (I), recovered from the infection (R), and those who died due to disease (D).

Given that intervention measures, applied in response to the emergence of COVID-19, are not the same for all population strata, we propose the use of multi-compartment version of standard SEIRDS model, named SEAIHRDS model. In this augmented classic deterministic model, the susceptible population was further stratified within the compartment S according to age and occupations. In addition, also two new compartments were added, asymptomatic cases of infection and a portion of the infected population who were hospitalized. To simulate the epidemic progression through different population strata-subgroups, we used appropriate, stratum-specific, model parameters and factors of effective contact reduction (anti-epidemic intervention measures - $\rho$), which were adapted to the relevant population groups. During the simulation, the effects of various levels of contact reduction, ranging from 20% to 75% were monitored, taking into account the realistic possibilities of maintaining a minimum work process, functioning of the society and feasibility of such measures.

Based on above assumption, the model of differential equations is formulated as follows:

$$\frac{dS_i(t)}{dt} = bN(t) - \sum_{j=1}^{n} \beta_{ij} I_j(t) S_i(t) - \sum_{j=1}^{n} \beta_{ij} \eta A_j(t) S_i(t) + \omega R_i(t) - m S_i(t)$$

$$\frac{dE_i(t)}{dt} = \sum_{j=1}^{n} \beta_{ij} I_j(t) S_i(t) + \sum_{j=1}^{n} \beta_{ij} \eta A_j(t) S_i(t) - (m + \mathfrak{f}) E_i(t)$$

$$\frac{dA_i(t)}{dt} = \mathfrak{f} \alpha E_i(t) - (\gamma + m) A_i(t)$$

$$\frac{dI_i(t)}{dt} = \mathfrak{f}(1 - \alpha) E_i(t) - \sum_{j=1}^{n} \sigma_{ij} I_j(t) - (r + m) I_i(t)$$

$$\frac{dH_i(t)}{dt} = \sum_{j=1}^{n} \sigma_{ij} I_j(t) - \sum_{j=1}^{n} \delta_{ij} H_i(t) - (\varepsilon + m) H_i(t)$$

$$\frac{dR_i(t)}{dt} = \sum_{j=1}^{n} r_{ij} I_j(t) + \sum_{j=1}^{n} \gamma_{ij} A_j(t) + \varepsilon H_i(t) - (m + \omega) R_i(t)$$

$$\frac{dD_i(t)}{dt} = \sum_{j=1}^{n} \delta_{ij} I_i(t).$$

In this model susceptible, exposed, infectious, recovered, deaths and total population are:

$S(t)=S_{ps}(t)+S_{es}(t)+S_{hs}(t)+S_{cs}(t)+S_{ua}(t)+S_{ea}(t)+S_p(t)$

$E(t)=E_{ps}(t)+E_{es}(t)+E_{hs}(t)+E_{cs}(t)+E_{ua}(t)+E_{ea}(t)+E_p(t)$

$A(t)=A_{ps}(t)+A_{es}(t)+A_{hs}(t)+A_{cs}(t)+A_{ua}(t)+A_{ea}(t)+A_p(t)$

$I(t)=I_{ps}(t)+I_{es}(t)+I_{hs}(t)+I_{cs}(t)+I_{ua}(t)+I_{ea}(t)+I_p(t)$

$H(t)=H_{ps}(t)+H_{es}(t)+H_{hs}(t)+H_{cs}(t)+H_{ua}(t)+H_{ea}(t)+H_p(t)$

$R(t)=R_{ps}(t)+R_{es}(t)+R_{hs}(t)+R_{cs}(t)+R_{ua}(t)+R_{ea}(t)+R_p(t)$

$D(t)=D_{ps}(t)+D_{es}(t)+D_{hs}(t)+D_{cs}(t)+D_{ua}(t)+D_{ea}(t)+D_p(t)$

$N(t)=S(t)+E(t)+I(t)+R(t)-D(t)$

$A_{t+1}=f\alpha E_t - (\gamma+m)A_t$

In order to simulate the possibility of controlling the epidemic with the help of vaccination, additional compartment to the model was added denoted with V(t), in which there are vaccinated persons who have successfully developed protective immunity after vaccination. The change of rate in this compartment per unit time is as follow:

$$\frac{dV_i(t)}{dt} = v_e \psi S_i(t) - mV_i - \omega V_i.$$

and compartment S(t) is slightly modified as follow:

$$\frac{dS_i(t)}{dt} = bN(t) - \sum_{j=1}^{n} \beta_{ij} I_j(t)\, S_i(t) + \omega R\_i\ (t) - mS_i(t) - v_e \psi S_i(t) + \omega V_i.$$

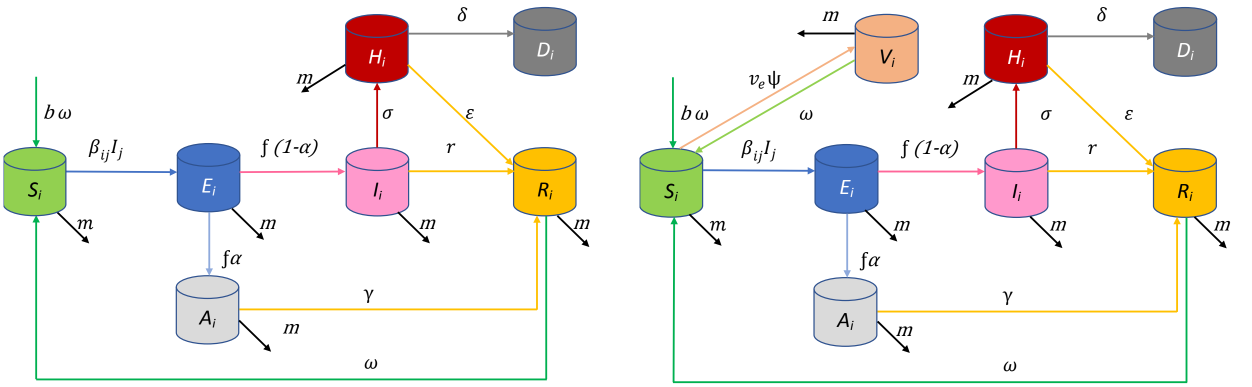


a) SEAIHRDS model with demography

b) SEAIHRDVS model with demography and vaccination

Fig. 1. Schematic representation of a mathematical model

#### 1.1. Model parameterisation

A summary of all model parameters is given in Tables 1, 2 and 3.

Table 1. Structure of different population strata in the Republic of Serbia [23]

| Stratum | Population | Percentage of total population |
|---|---|---|
| Younger than 7 years | 356,377 | 5.10% |
| Elementary school | 550,527 | 7.88% |
| Secondary school | 249,455 | 3.57% |
| Students | 241,698 | 3.46% |
| Employed | 2,197,065 | 31.46% |
| Pensioners | 1,715,152 | 24.56% |
| Others | 1,672,330 | 23.95% |

Table 2. Age structure of the population of the Republic of Serbia and expected percentage of hospitalized patients, patients in intensive care, and death rate caused by COVID-19.

| Population age groups | *Population | Percentage of total population | **Expected % of hospitalized patients (σ) | ***Expected % of patients whose treatment requires intensive care | ****Infection fatality rate IFR (male/female) |
|---|---|---|---|---|---|
| 0-9 | 458,199 | 6.56% | 0.00% | 5.00% | 0.04%;0.01% |
| 10-19 | 445,481 | 6.38% | 0.04% | 5.00% | 0.00%;0.02% |
| 20-29 | 1,028,226 | 14.73% | 1.04% | 5.00% | 0.00%;0.01% |
| 30-39 | 951,615 | 13.63% | 3.43% | 5.00% | 0.00%;0.05% |
| 40-49 | 968,854 | 13.88% | 4.25% | 6.30% | 0.08%;0.04% |
| 50-59 | 963,229 | 13.79% | 8.16% | 12.20% | 0.33%;0.20% |
| 60-69 | 815,244 | 11.68% | 11.80% | 27.40% | 1.62%;0.62% |
| 70-79 | 696,045 | 9.97% | 16.60% | 43.20% | 6.11%;2.68% |
| 80- | 655,711 | 9.39% | 18.40% | 70.90% | 16.40%;6.49% |

*[ 26], **[30], ***[8], ***[31]

Table 3. SEAIHRD model parameters

| Input parameters | Mark | Value | Source |
|---|---|---|---|
| Population | $N_{t0}$ | 6,982,604 | [26] |
| Initial number of cases | $It_0$ | 1 | Fixed number |
| Initially immune | $I_{mm0}$ | 0 | Fixed number |
| Basic reproduction number | $R_0$ | 2.46 (3.1) | [32] |
| Effective contact rate | $C_e$ | 0.38 | Estimated |
| *Per capita* contact rate | $\beta$ | 0.0000000378 | Estimated |
| Daily infection rate (transfer E→I) | $f$ | 0.294118 | Estimated |
| Recovery rate of symptomatic cases | $r$ | 0.107527 | Estimated |
| Daily rate of waning of immunity | $\omega$ | 0.002739726 | Estimated |
| *Per capita* birth rate | $b$ | 0.000025205 | [26] |
| *Per capita* death rate unrelated to COVID-19 | $m$ | 0.000036006 | Estimated |
| Life expectancy in years | $L$ | 76.09 | [27] |
| Duration of latent infection in days | $f^{-1}$ | 3.5 | [28] |
| Duration of infectious period in days (clinical cases) | $T_R$ | 9.3 | [28] |
| Duration of immunity in days | $I_{mm}$ | 365.00 | Assumed |
| Incubation period in days | *Inc* | 5.8 | [33] |
| Time period (day) | *dt* | 1.00 | - |
| Average treatment duration in hospital | *days* | 15,9 | [34] |
| Average time spent in intensive care | *days* | 27 | [34] |
| Recovery rate of hospitalized cases | $\varepsilon$ | 0.062893 | Estimated |
| Average times taken from onset of symptoms to death | *days* | 17 | [35] |
| Infectious period for asymptomatic cases | *days* | 7.25 | [28, 36] |
| Recovery rate of asymptomatic cases | $\gamma$ | 0.137931 | Estimated |
| Expected percentage of asymptomatic cases | - | 30% | [37] |
| Infectiousness of asymptomatic cases in relation to symptomatic cases | $\eta$ | 75% | [37] |

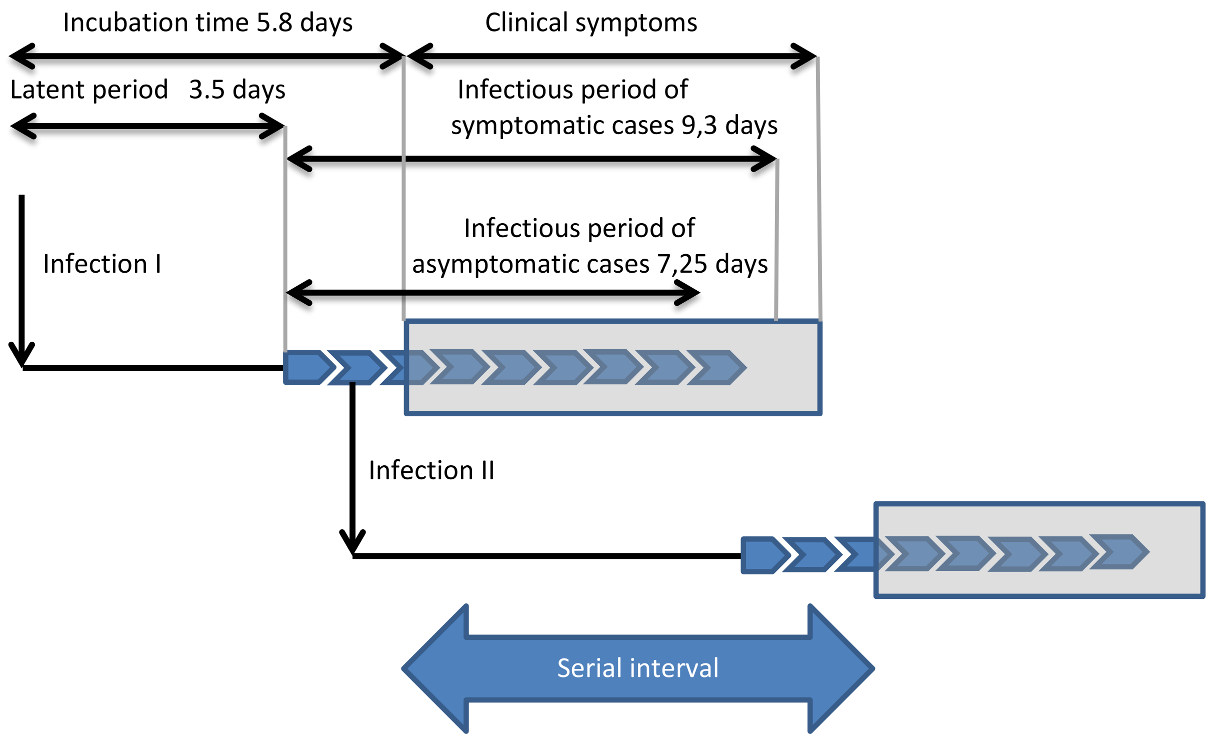


Fig.2. Summary of definitions of incubation period, latent period (pre-infectious) and infectious period for COVID-19.

#### 1.2. Model validation

Once the model has been created, there is a need to compute objective metrics to evaluate whether the model generated good predicted values with regard to the variable under study.

During the validation process, the historical data of the actual epidemic of COVID-19 in Serbia were compared with the data obtained by the SEAIHRDS model. For validation purposes, the current epidemic of COVID-19 was simulated (scenario marked as SC0_SER_COVID-19), along with actual anti-epidemic measures, and then the results of simulation was compared with cumulative number of deaths officially recorded.

In the first phase of validation, with the help of the model, the conditions under which the COVID-19 epidemic spread were mimic, as well as the measures of interventions that were taken in order to stop the epidemic. The obtained data were then analysed using statistical tests. We used quality measures as follows:

1) Mean absolute error, given by equation

$$MAE(y,\hat{y}) = \frac{1}{N}\sum_{n=1}^{N} \left| y(t) - \hat{y}(t) \right|$$

2) Mean squared error, given by equation

$$MSE(y,\hat{y}) = \frac{1}{N}\sum_{n=1}^{N} (y(t) - \hat{y}(t))^2$$

3) Root mean square error, given by equation

$$RMSE(y,\hat{y}) = \sqrt{\frac{1}{N}\sum_{n=1}^{N} (y(t) - \hat{y}(t))^2}$$

4) Normalized mean average error, given by equation

$$NormMAE(y,\hat{y}) = \frac{MAE(y,\hat{y})}{\bar{y}}$$

5) We calculate the maximum deviation between the main prediction line and SC0_SER_COVID-19 using a 95% confidence level. The equation of this measure is

$$MaxDev(y,\hat{y}) = \frac{\left| y(t) - \hat{y}(t) \right|}{y(t)}$$

Pearson's R and coefficient of determination, $R^2$, was used to check goodness of fit of SEAIHRD model with COVID-19 data recorded during the real epidemic.

The following formula was used to calculate the coefficient of determination

$$R^2 = \frac{\sum_{n+1}^{N} (\hat{y} - \bar{y})^2}{\sum (y - \bar{y})^2}$$

and for Pearson's R,

$$R = \frac{\sum_{n=1}^{N} (x - \bar{x})(y - \bar{y})}{\sqrt{\sum_{n=1}^{N} (x - \bar{x})^2 \sum_{n=1}^{N} (y - \bar{y})^2}} \quad .$$

### 2. Results

#### 2.1. Predicting the number of sick, hospitalized patients and deaths caused by COVID-19 in Serbia

Table 4 and Fig. 2 show the summarized simulation results of 5 different scenarios. Scenario number 1 represents a prediction of the possible outcome of the spread of the epidemic in the absence of any intervention measures. The other 4 scenarios represent the possible outcomes of the epidemic depending on the intensity of the applied measures. Fig. 1 and 2 show the long-term predictions of the possible dynamics of the COVID-19 epidemic in Serbia in the absence of any intervention measures.

Fig. 1, panels a) and b) show daily variations in the number of susceptible, latently infected, infected and recovered patients, at basic reproduction numbers of $R_o$=2.46 and $R_o$=3, respectively. Panels c) and d) of the same figure show daily fluctuations in susceptible, recovered and net reproduction rates $R_n$ for $R_o$=2.46 and $R_o$=3, respectively. Panels e) and f) of Fig. 1 show daily variations in $R_{n,}$ true and apparent disease incidences at basic reproduction numbers of $R_o$=2.46 and $R_o$=3.

Fig. 2, panels a) and b) show a prediction of necessary hospital capacities. Panels c) and d) of Fig. 2 show predicted numbers of sick and dead due to COVID-19 at $R_o$=2.46 and $R_o$=3 and age structure of hospitalized patients and deaths.

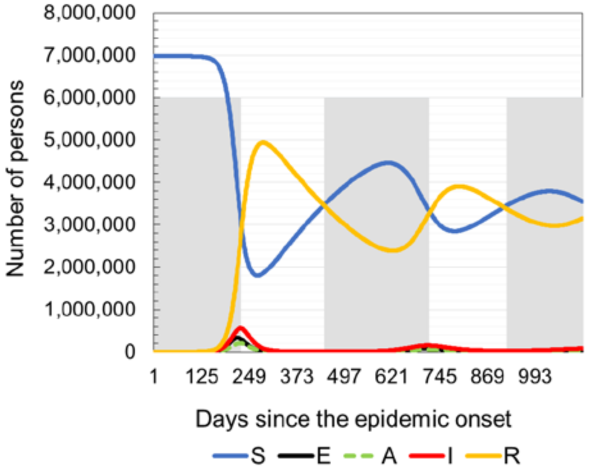


a) Distribution of the total number of the susceptible, latently infected, asymptomatic infectious individuals, infected and recovered on a daily basis since the epidemic onset ($R_o$=2.46)

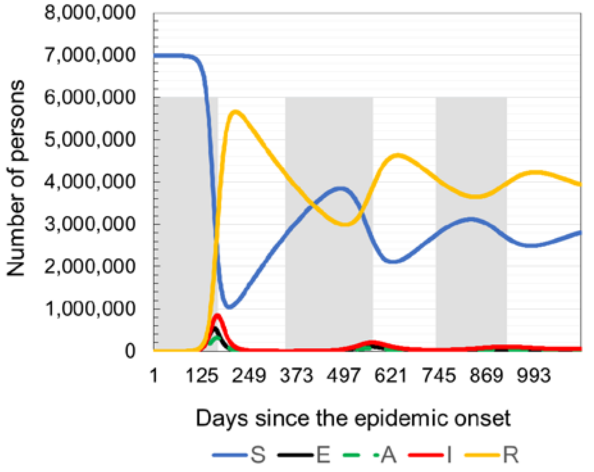


b) Distribution of the total number of the susceptible, latently infected, asymptomatic infectious individuals, infected and recovered on a daily basis since the epidemic onset ($R_o$=3.1)

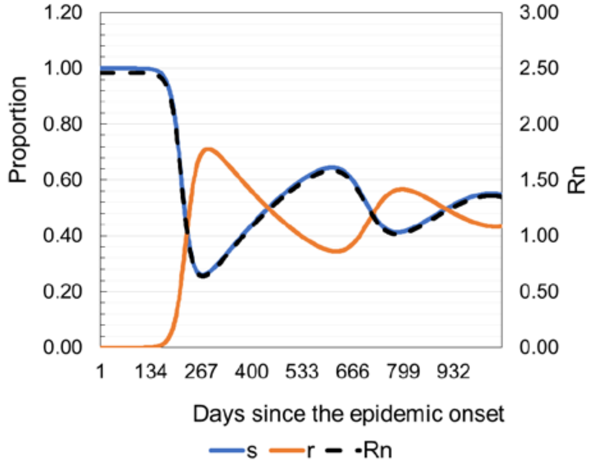


c) Daily fluctuations of the susceptible, recovered and net disease transmission rates ($R_o$=2.46)

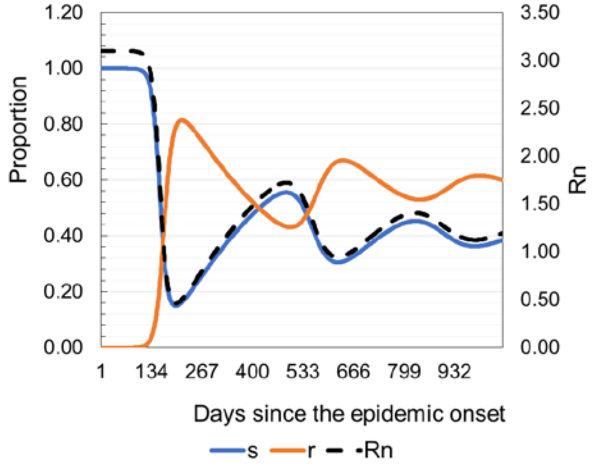


d) Daily fluctuations of the susceptible, recovered and net disease transmission rates ($R_o$=3.1)

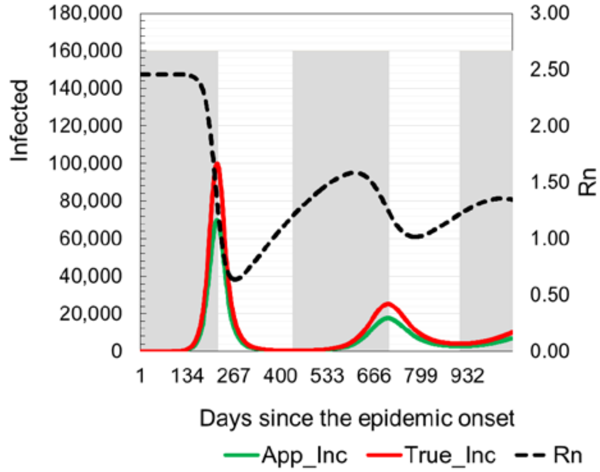


e) Daily fluctuations of apparent incidence, true incidence and net disease transmission rates ($R_o$=2.46)

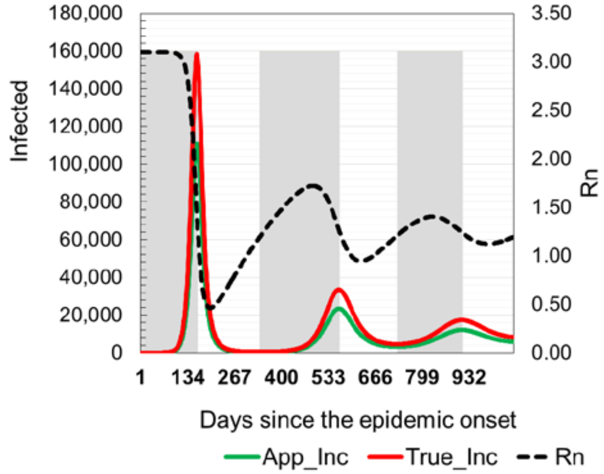


f) Daily fluctuations of apparent incidence, true incidence and net disease transmission rates ($R_o$=2.46)

Fig.1. Model prediction of latently infected, diseased, recovered and daily fluctuations of $R_n$.

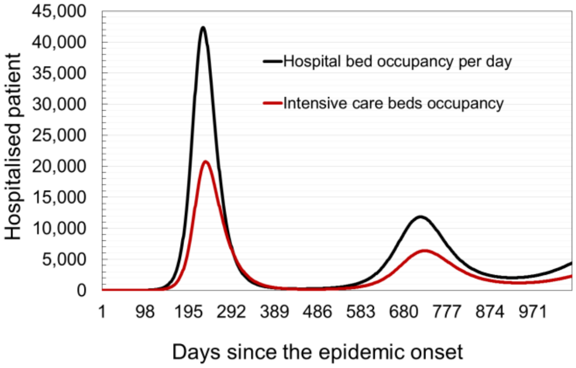


a) Necessary hospital capacities ($R_o$=2.46)

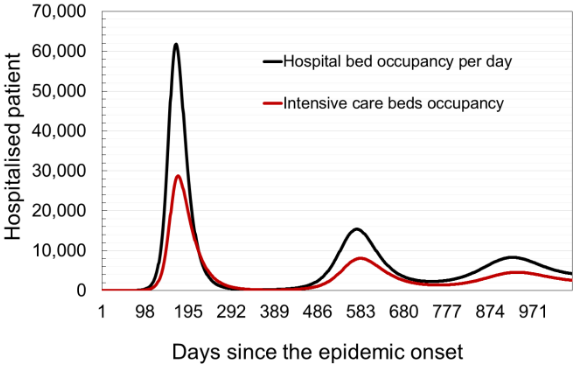


b) Necessary hospital capacities ($R_o$=3.1)

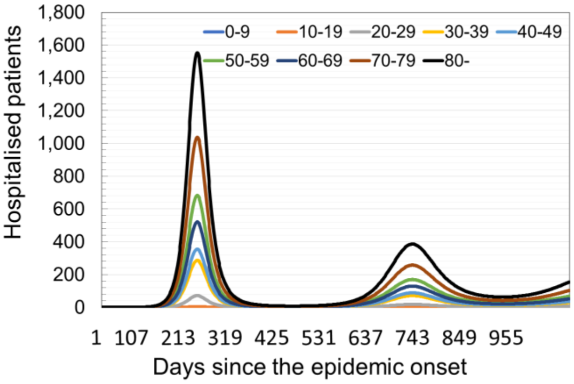


c) Age structure of hospitalized patients ($R_o$=2.46)

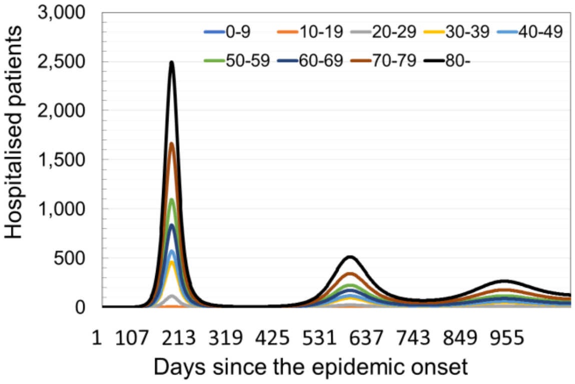


d) Age structure of hospitalized patients ($R_o$=3.1)

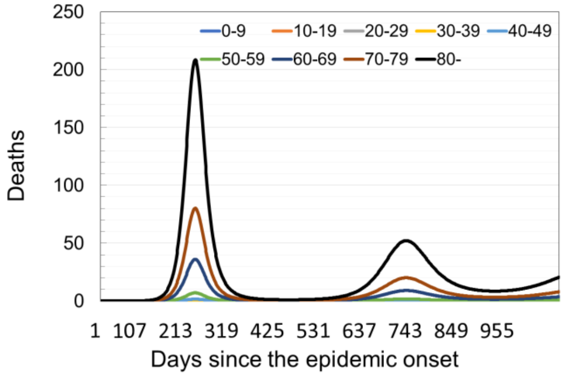


e) Expected age structure of the dead ($R_o$=2.46)

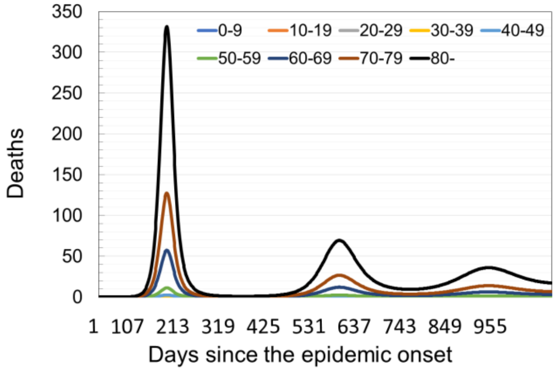


f) Expected age structure of the dead (Ro=3.1)

Fig. 2. Model prediction of required hospital capacities under the assumption of different intervention measures.

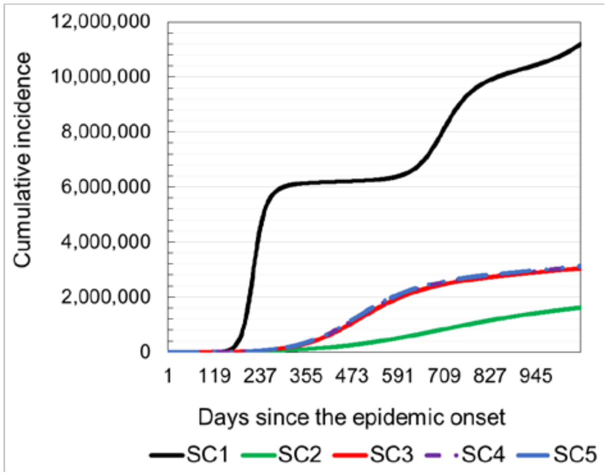


a) Comparative overview of cumulative incidences (CI). Results obtained from simulated scenarios 1-5 ($R_o$=2.46)

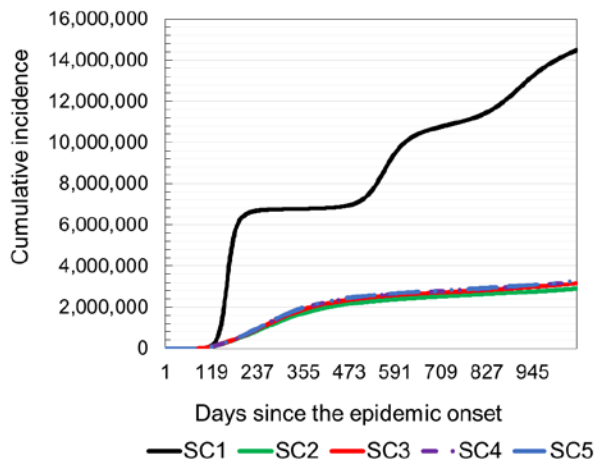


b) Comparative overview of cumulative incidences (CI). Results obtained from simulated scenarios 1-5 ($R_o$=3.1)

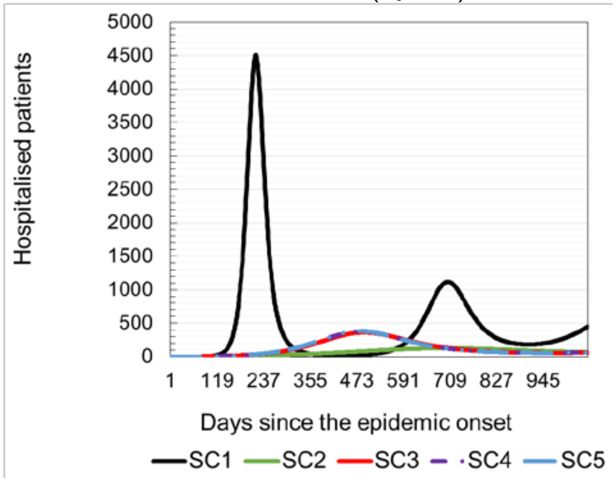


c) Comparative overview of hospitalized patients on a daily basis. Results obtained from simulated scenarios 1-5 ($R_o$=2.46)

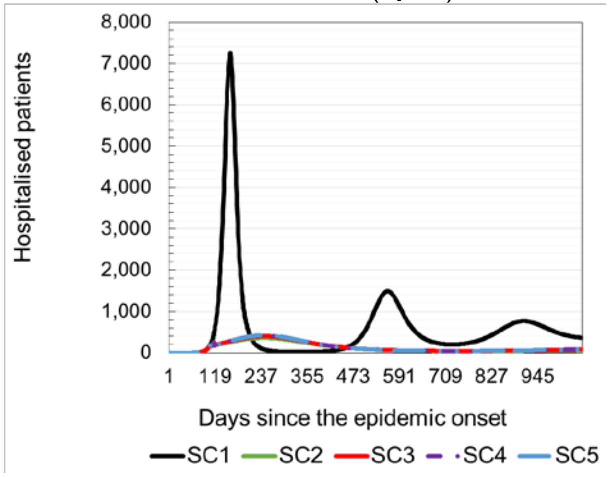


d) Comparative overview of hospitalized patients on a daily basis. Results obtained from simulated scenarios 1-5 ($R_o$=3.1)

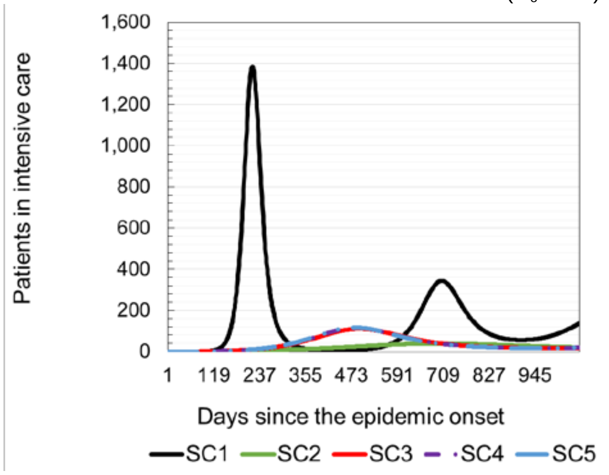


e) Comparative overview of expected number of patients in intensive care on a daily basis. Results obtained from simulated scenarios 1-5 with $R_o$=2.46

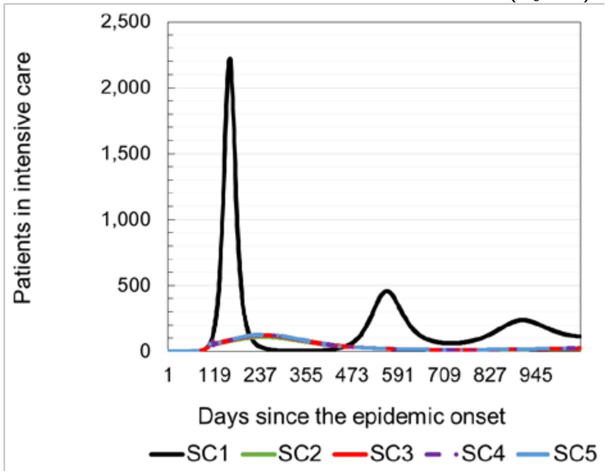


f) Comparative overview of expected number of patients in intensive care on a daily basis. Results obtained from simulated scenarios 1-5 with $R_o$=3.1

Fig.3. Model prediction of expected number of hospitalized patient and patient in intensive care units.

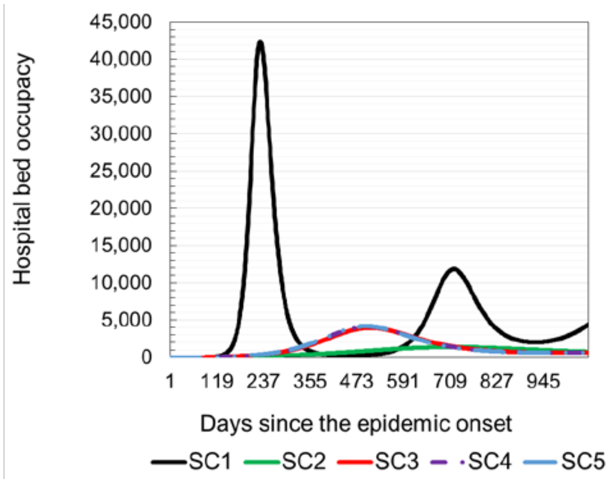


a) Comparative overview of hospital bed occupancy ($R_o$=2.46)

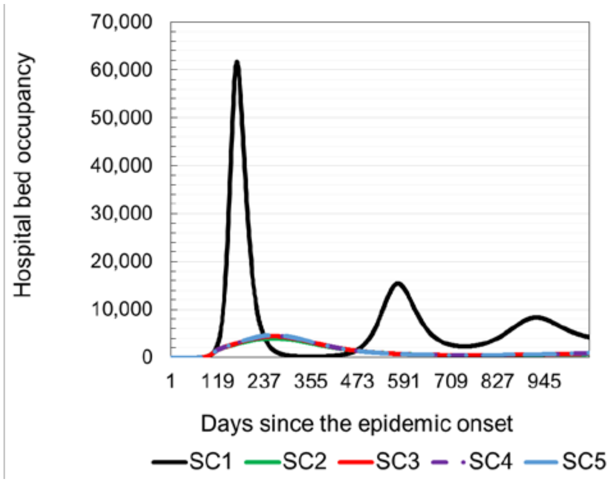


b Comparative overview of hospital bed occupancy ($R_o$=3.1)

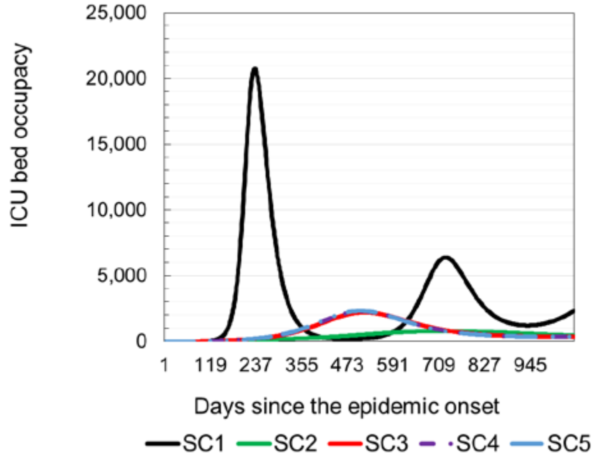


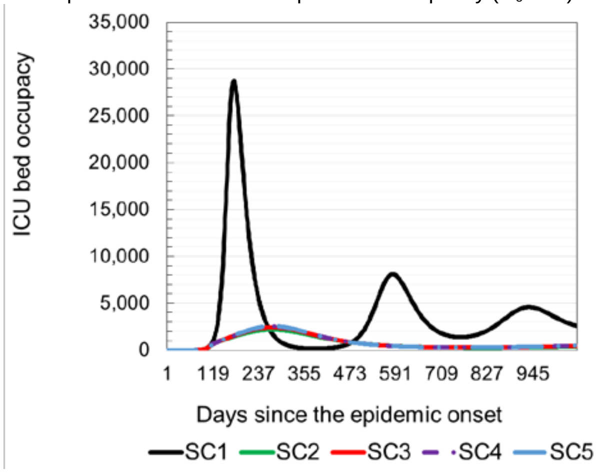


c) Comparative overview of ICU bed occupancy (Ro=2.46)

d) Comparative overview of ICU bed occupancy (Ro=3.1)

Fig. 4. Model prediction of required hospital capacities needed to treat patients with COVID-19.

Table 4. Results of different simulated scenarios (Ro = 2.46 and Ro = 3.1). The data refers to the period of 365 days from epidemic onset.

| Scenario mark | SC1 | | SC2 | | SC3 | | SC4 | | SC5 | |
|---|---|---|---|---|---|---|---|---|---|---|
| Basic reproductive number | $R_o$=2.64 | $R_o$=3.1 | $R_o$=2.64 | $R_o$=3.1 | $R_o$=2.64 | $R_o$=3.1 | $R_o$=2.64 | $R_o$=3.1 | $R_o$=2.64 | $R_o$=3.1 |
| Cumulative incidence (CI) | 6,229,144 | 7,133,221 | 308,581 | 2,219,251 | 1,286,227 | 2,419,079 | 1,375,416 | 2,489,197 | 1,408,262 | 2,514,936 |
| Apparent CI | 4,360,401 | 4,993,254 | 216,007 | 1,553,476 | 900,359 | 1,693,355 | 962,791 | 1,742,438 | 985,783 | 1,760,456 |
| Overall hospitalized | 278,781 | 320,567 | 13,970 | 98,337 | 56,631 | 105,930 | 60,319 | 108,554 | 61,685 | 109,538 |
| Overall in intensive care | 85,633 | 98,333 | 4,260 | 29,753 | 17,085 | 32,058 | 18,202 | 32,855 | 18,612 | 33,147 |
| Overall deaths | 20,894 | 23,951 | 1,031 | 7,194 | 4,113 | 7,754 | 4,383 | 7,948 | 4,483 | 8,018 |

### 2.2. Predicting the number of sick, hospitalized patients and deaths caused by COVID-19 in Serbia in the case of the application of current anti-epidemic measures in simulation

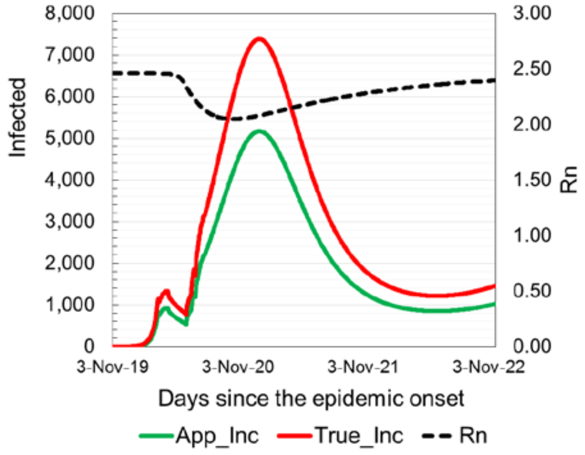


a) Daily fluctuations of apparent incidence, true incidence and net disease transmission rates ($R_o$=2.46)

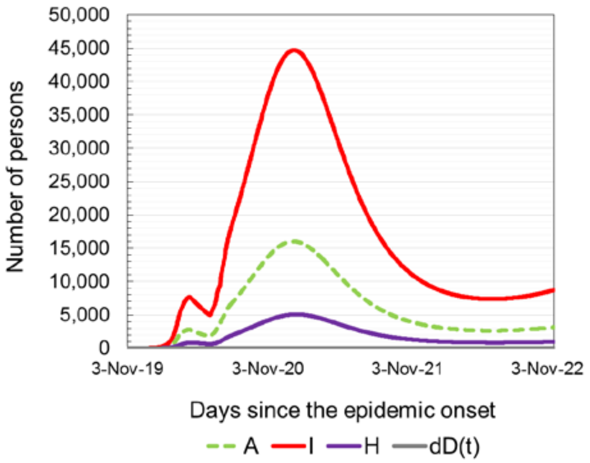


b) Distribution of the total number of asymptomatic cases, infected, hospitalized patients and deaths on a daily basis since the epidemic onset ($R_o$=2.46)

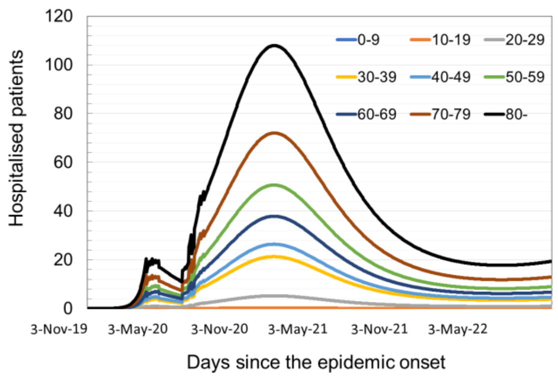


c) Age structure of hospitalized patients ($R_o$=2.46)

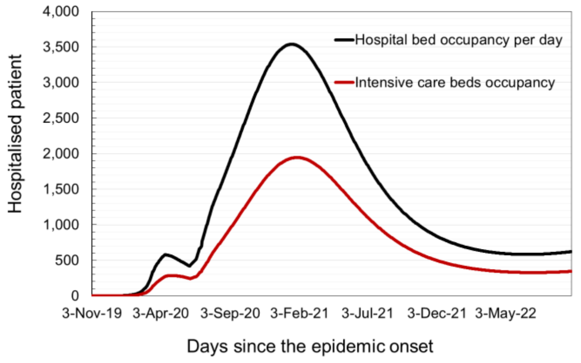


d) Comparative overview of hospitalized patients on a daily basis. Results obtained from simulated scenarios 1-5 ($R_o$=3.1)

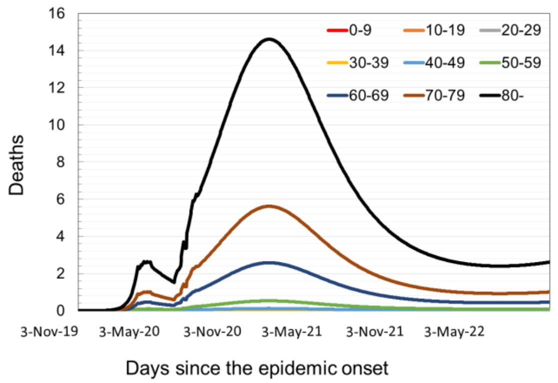


e) Expected age structure of the dead (Ro=2.46)

- Cumulative incidence: 1,611,871
- Apparent incidence: 1,128,309
- Total hospitalized patients: 70,945
- Total in ICU: 21,380
- Total recovered: 1,606,715
- Total deceased: 5,155

f) Overview of expected number of infected, hospitalized and patients in intensive care units and deceased in the first year of epidemic ($R_o$=2.46).

Fig. 5. Predictions of possible epidemic dynamics in case of application of current anti-epidemic measures

#### 2.3. Predicting the number of sick, hospitalized patients and deaths caused by COVID-19 in Serbia in the case of the application of vaccination strategy in simulation

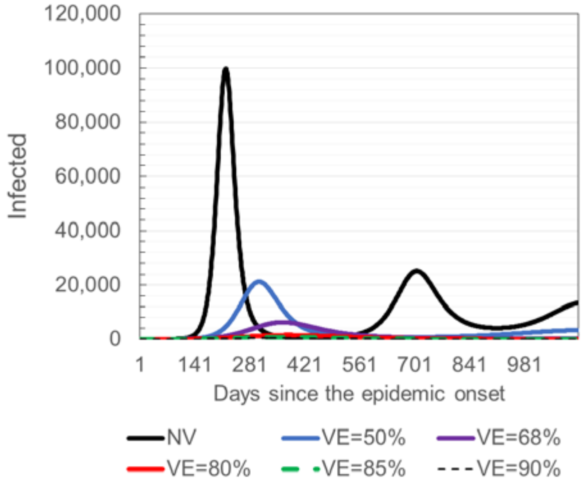


a) Comparative overview of daily incidences ($R_o$=2.46).

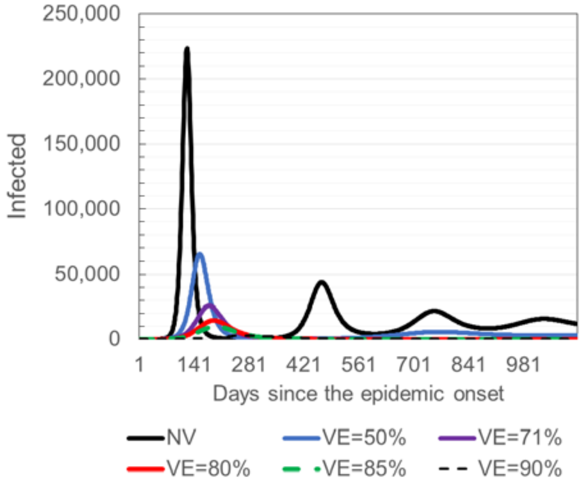


b) Comparative overview of daily incidences ($R_o$=3.1).

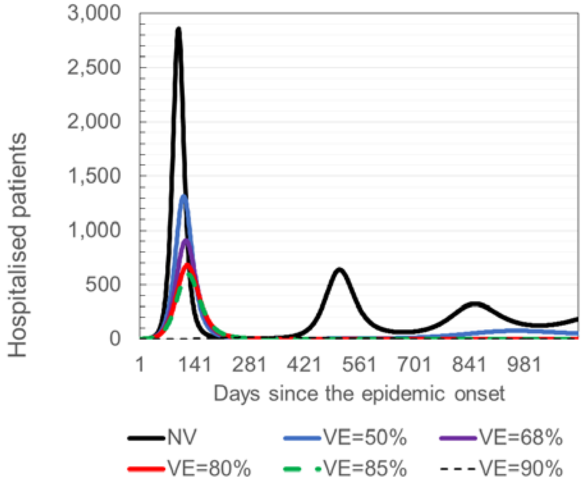


c) Comparative overview of hospitalized patients ($R_o$=2.46).

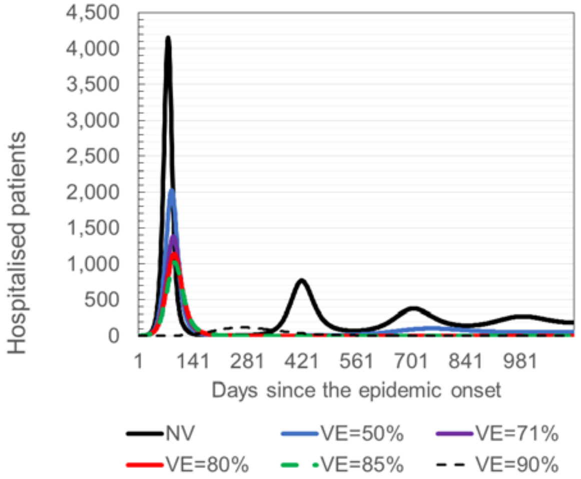


d) c) Comparative overview of hospitalized patients ($R_o$=3.1).

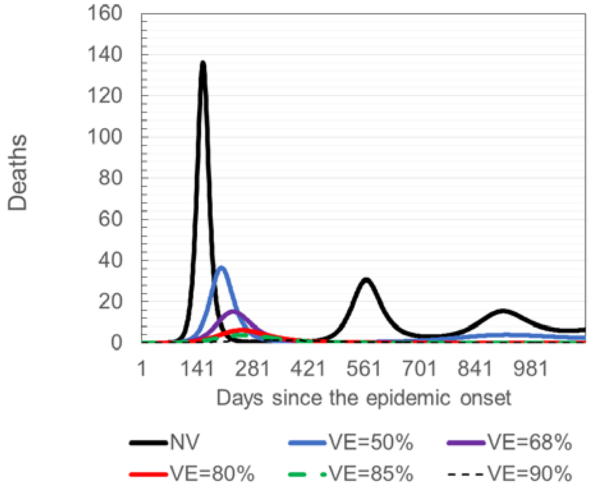


e) Comparative overview of deaths (Ro=2.46)

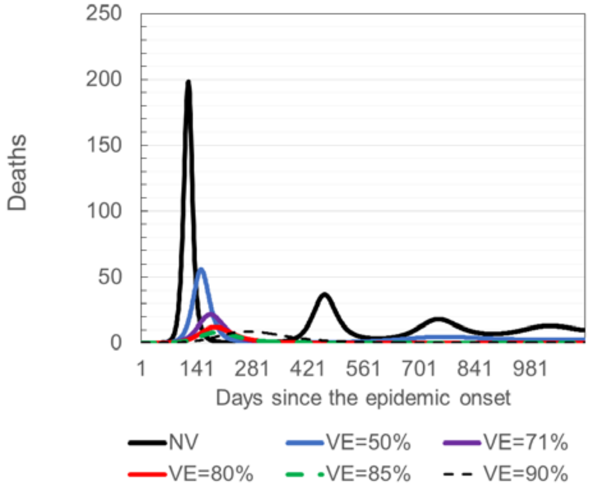


f) Comparative overview of deaths (Ro=3.1)

Fig. 6. Results of simulated COVID-19 control based solely on vaccination, scenarios 6-9 (Ro=2.46; Ro=3.1).

Depending on the efficacy of the potential vaccine, the required vaccination coverage should be 87% ($v_e$ =68%), 74.19% ($v_e$ =80%), 69.82% ($v_e$=85%), 65.94% ($v_e$=90%) and 95.41% ($v_e$=71%), 84.68% ($v_e$=80%), 79.70% ($v_e$=85%), 75.27% ($v_e$=90%) for $R_o$=2.46 and $R_o$=3.1, respectively.

Table 6. Results of simulated scenarios 6-10 (Ro = 2.46 and Ro = 3.1). The data refers to the period of 1 year of epidemic.

| Scenario mark | SC6 ($V_e$=50%) | | SC7 ($V_e$=68%) | | SC8 ($V_e$=80%) | | SC9 ($V_e$=85%) | | SC10 ($V_e$=90%) | |
|---|---|---|---|---|---|---|---|---|---|---|
| Basic reproductive number | $R_o$=2.64 | $R_o$=3.1 | $R_o$=2.64 | $R_o$=3.1 | $R_o$=2.64 | $R_o$=3.1 | $R_o$=2.64 | $R_o$=3.1 | $R_o$=2.64 | $R_o$=3.1 |
| Cumulative incidence (CI) | 2,840,334 | 3,642,845 | 1,130,361 | 2,277,004 | 315,832 | 1,260,081 | 169,680 | 859,450 | 91,870 | 531,339 |
| Apparent CI | 1,988,234 | 2,549,992 | 791,253 | 1,593,903 | 221,082 | 882,057 | 118,776 | 601,615 | 64,309 | 371,937 |
| Overall hospitalized | 126,962 | 162,932 | 50,776 | 101,760 | 14,210 | 56,323 | 7,624 | 38,427 | 4,122 | 23,761 |
| Overall in intensive care | 38,976 | 50,047 | 15,550 | 31,252 | 4,349 | 17,291 | 2,335 | 11,795 | 1,263 | 7,293 |
| Overall deaths | 9,502 | 12,211 | 3,777 | 7,624 | 1,056 | 4,216 | 567 | 2,875 | 307 | 1,777 |

### 2.4. Results of the model validation

Table 7. Regression model

| date | x ($D_{\_observed}$) | y ($_{(D\_SEAIHRD)}$) | ŷ=ax+b | *Regression Statistics* | |
|---|---|---|---|---|---|
| 1-Dec-20 | 1,652.00 | 2,802.90 | 2,667.83 | Multiple R | 0.981678289 |
| 2-Dec-20 | 1,704.00 | 2,825.59 | 2,698.98 | R Square | 0.963692263 |
| 3-Dec-20 | 1,765.00 | 2,848.32 | 2,735.53 | Adjusted R Square | 0.963124954 |
| 4-Dec-20 | 1,834.00 | 2,871.11 | 2,776.86 | Standard Error | 86.24143528 |
| 5-Dec-20 | 1,891.00 | 2,893.95 | 2,811.01 | Observations | 66 |
| 6-Dec-20 | 1,949.00 | 2,916.84 | 2,845.75 | | |
| 7-Dec-20 | 2,005.00 | 2,939.77 | 2,879.30 | | |
| 8-Dec-20 | 2,062.00 | 2,962.75 | 2,913.44 | | |
| 9-Dec-20 | 2,116.00 | 2,985.77 | 2,945.79 | | |
| 10-Dec-20 | 2,172.00 | 3,008.83 | 2,979.34 | | |
| 11-Dec-20 | 2,227.00 | 3,031.93 | 3,012.29 | | |
| 12-Dec-20 | 2,275.00 | 3,055.08 | 3,041.04 | | |
| 13-Dec-20 | 2,331.00 | 3,078.25 | 3,074.59 | | |
| 14-Dec-20 | 2,380.00 | 3,101.47 | 3,103.94 | | |
| 15-Dec-20 | 2,433.00 | 3,124.71 | 3,135.69 | | |
| 16-Dec-20 | 2,482.00 | 3,147.99 | 3,165.04 | | |
| 17-Dec-20 | 2,529.00 | 3,171.30 | 3,193.20 | | |
| 18-Dec-20 | 2,580.00 | 3,194.64 | 3,223.75 | | |
| 19-Dec-20 | 2,632.00 | 3,218.00 | 3,254.90 | | |
| 20-Dec-20 | 2,686.00 | 3,241.39 | 3,287.25 | | |
| 21-Dec-20 | 2,733.00 | 3,264.81 | 3,315.41 | | |
| 22-Dec-20 | 2,782.00 | 3,288.24 | 3,344.76 | | |
| 23-Dec-20 | 2,833.00 | 3,311.70 | 3,375.31 | | |
| 24-Dec-20 | 2,882.00 | 3,335.17 | 3,404.66 | | |
| 25-Dec-20 | 2,931.00 | 3,358.66 | 3,434.02 | | |
| 26-Dec-20 | 2,983.00 | 3,382.17 | 3,465.17 | | |
| 27-Dec-20 | 3,030.00 | 3,405.69 | 3,493.32 | | |
| 28-Dec-20 | 3,073.00 | 3,429.23 | 3,519.08 | | |
| 29-Dec-20 | 3,119.00 | 3,452.77 | 3,546.64 | | |
| 30-Dec-20 | 3,163.00 | 3,476.32 | 3,573.00 | | |
| 31-Dec-20 | 3,211.00 | 3,499.88 | 3,601.75 | | |
| 1-Jan-21 | 3,250.00 | 3,523.45 | 3,625.12 | | |
| 2-Jan-21 | 3,288.00 | 3,547.02 | 3,647.88 | | |
| 3-Jan-21 | 3,325.00 | 3,570.59 | 3,670.04 | | |
| 4-Jan-21 | 3,364.00 | 3,594.16 | 3,693.41 | | |
| 5-Jan-21 | 3,405.00 | 3,617.73 | 3,717.97 | | |
| 6-Jan-21 | 3,444.00 | 3,641.30 | 3,741.33 | | |

| | | | |
|---|---|---|---|
| 7-Jan-21 | 3,479.00 | 3,664.86 | 3,762.30 |
| 8-Jan-21 | 3,513.00 | 3,688.42 | 3,782.67 |
| 9-Jan-21 | 3,548.00 | 3,711.97 | 3,803.63 |
| 10-Jan-21 | 3,582.00 | 3,735.51 | 3,824.00 |
| 11-Jan-21 | 3,610.00 | 3,759.04 | 3,840.77 |
| 12-Jan-21 | 3,639.00 | 3,782.56 | 3,858.15 |
| 13-Jan-21 | 3,664.00 | 3,806.07 | 3,873.12 |
| 14-Jan-21 | 3,687.00 | 3,829.55 | 3,886.90 |
| 15-Jan-21 | 3,708.00 | 3,853.03 | 3,899.48 |
| 16-Jan-21 | 3,730.00 | 3,876.48 | 3,912.66 |
| 17-Jan-21 | 3,750.00 | 3,899.91 | 3,924.64 |
| 18-Jan-21 | 3,771.00 | 3,923.33 | 3,937.22 |
| 19-Jan-21 | 3,791.00 | 3,946.71 | 3,949.20 |
| 20-Jan-21 | 3,810.00 | 3,970.08 | 3,960.58 |
| 21-Jan-21 | 3,830.00 | 3,993.42 | 3,972.56 |
| 22-Jan-21 | 3,849.00 | 4,016.73 | 3,983.95 |
| 23-Jan-21 | 3,868.00 | 4,040.01 | 3,995.33 |
| 24-Jan-21 | 3,886.00 | 4,063.26 | 4,006.11 |
| 25-Jan-21 | 3,905.00 | 4,086.48 | 4,017.49 |
| 26-Jan-21 | 3,924.00 | 4,109.66 | 4,028.88 |
| 27-Jan-21 | 3,944.00 | 4,132.81 | 4,040.86 |
| 28-Jan-21 | 3,965.00 | 4,155.93 | 4,053.44 |
| 29-Jan-21 | 3,983.00 | 4,179.01 | 4,064.22 |
| 30-Jan-21 | 4,000.00 | 4,202.05 | 4,074.40 |
| 31-Jan-21 | 4,020.00 | 4,225.04 | 4,086.38 |
| 1-Feb-21 | 4,038.00 | 4,248.00 | 4,097.17 |
| 2-Feb-21 | 4,056.00 | 4,270.91 | 4,107.95 |
| 3-Feb-21 | 4,071.00 | 4,293.78 | 4,116.94 |
| 4-Feb-21 | 4,085.00 | 4,316.61 | 4,125.32 |

ANOVA

| | df | SS | MS | F | Significance F |
|---|---|---|---|---|---|
| Regression | 1 | 12634297.88 | 12634297.88 | 1698.709676 | 8.41222E-48 |
| Residual | 64 | 476005.4502 | 7437.58516 | | |
| Total | 65 | 13110303.33 | | | |

| | Coefficients | Standard Error | t Stat | P-value | Lower 95% | Upper 95% | Lower 95.0% | Upper 95.0% |
|---|---|---|---|---|---|---|---|---|
| Intercept | 1678.20184 | 46.85425976 | 35.817487 | 4.63985E-44 | 1584.599696 | 1771.80398 | 1584.59969 | 1771.80398 |
| x (D_observed) | 0.599050366 | 0.014534623 | 41.21540581 | 8.41222E-48 | 0.570014119 | 0.62808661 | 0.57001411 | 0.62808661 |

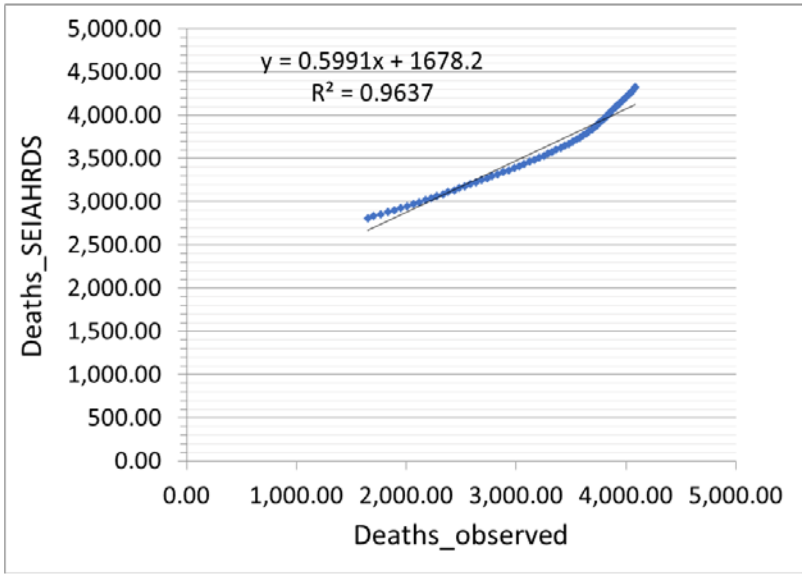


Fig. 7. Scatter diagram: deaths_observed vs deaths_predicted by SEAIHRDS model

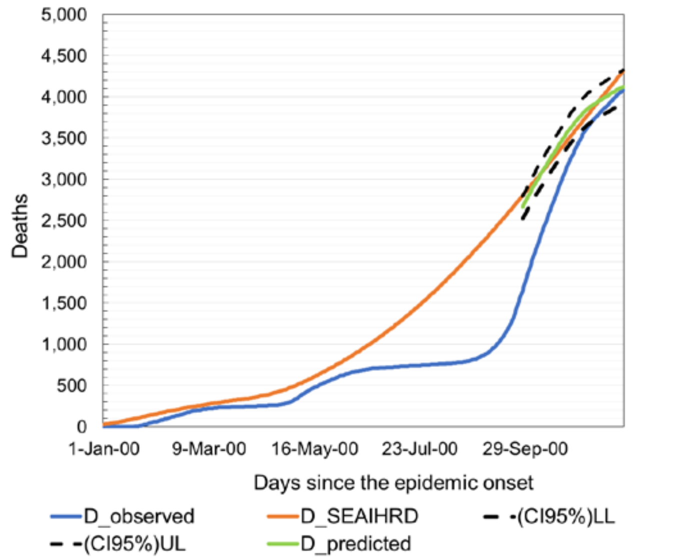


Fig. 8. The observed number of deceased individuals (blue), number of deceased individuals modeled with SEAIHRD model (orange), and predicted number of deceased individuals (green) by model and corrected with real data (95% confidence interval between dotted lines).

Table 11. Measures of the prediction quality

| | MAE | %Error | MSE | RMSE | Normalized MAE | Normalized MSE | Max Deviation |
|---|---|---|---|---|---|---|---|
| Deceased | 73 | 2.04% | 7,212.20 | 84.92 | 2.05% | 0.06% | 4.82% |
